# Countering the potential re-emergence of a deadly infectious disease - information warfare, identifying strategic threats, launching countermeasures

**DOI:** 10.1101/2020.10.13.20211680

**Authors:** Rex N. Ali, Harvey Rubin, Saswati Sarkar

**Author notes:** Email addresses (Rex N. Ali), (Harvey Rubin), (Saswati Sarkar).

## Abstract

Eradicated infectious diseases like smallpox can re-emerge through accident or designs of bioterrorists, and perpetrate heavy casualties. Currently, only a small percentage of the populace is vaccinated, and their protection is likely to have waned. Most therefore are susceptible today. And when the disease re-emerges the susceptible individuals may be manipulated by disinformation on Social Media to refuse vaccines. Thus, a combination of countermeasures consisting of antiviral drugs and vaccines and a range of policies for their application need to be investigated. Opinions as to receptivity of vaccines evolve with time through social exchanges over networks that overlap with but are not identical to the disease propagation networks. These couple the spread of the biological and information contagion and necessitate a joint investigation of the two. Towards these, we develop a computationally tractable metapopulation epidemiological model that captures the joint spatio-temporal evolution of smallpox and opinion dynamics. The computations based on the model show that opinion dynamics has a substantial impact on the fatality count. Towards understanding how perpetrators are likely to seed the infection we identify a) the initial distribution of infected individuals that maximize the overall fatality count regardless of mobility patterns, and b) which habitation structures are more vulnerable to outbreaks. We assess the relative efficacy of different countermeasures and conclude that a combination of vaccines and drugs minimizes the fatalities, and by itself, for smallpox, drugs reduce fatalities more than the vaccine. Accordingly, we assess the efficacies of three separate policies for administering the drugs and identify the best among them for various parameter combinations. When the availability of the drug is finite, we show that increase in its supply substantially reduces the overall fatality. Our findings lead to policy recommendations for public health and urban design authorities towards thwarting smallpox and other infectious disease outbreaks.

## Introduction

The devastative potential of a sudden outbreak of an infectious disease is self-evident in this time of a pandemic. The havoc caused by the COVID-19 outbreak may not only be replicated but also amplified should an eradicated infectious disease re-emerge. While naturally occurring smallpox was eradicated in the 1970s through decades of a global vaccination campaign, it can re-emerge under various scenarios [14, 15, 20, 24]. Variola virus (smallpox virus) is a category A bioterrorism agent [2] and stocks of the virus are known to officially exist in two high-security biosafety level 4 laboratories in the United States (Centers for Disease Control and Prevention) and Russia (VECTOR Institute) and potentially elsewhere too [14, 15, 20, 24]. For instance, in 2014, scientists at the National Institute of Health (NIH) discovered half-dozen forgotten vials of smallpox in a storage room on its campus in Bethesda, Maryland [5, 30]. In addition, a virus similar to Variola, horsepox, has recently been synthesized from genetic pieces ordered in the mail [28], and smallpox may be recreated using similar techniques.

If smallpox does re-emerge, vaccine hesitancy - that is, refusing immunization on non-medical grounds (e.g., religious and philosophical beliefs, safety concern, disinformation) - can thwart attempts to proactively prevent it. Anti-vaccination movements also known as “Antivaxxers” have routinely propagated disinformation regarding immunization for various diseases e.g. that measles, mumps, and rubella (MMR) vaccine causes autism [8, 29]. A recent survey has shown that respectively less than 25% and 50% individuals will opt for a COVID-19 vaccine within three months and a year of its launch [10]. Vaccine hesitancy caused a spike in measles outbreak in 2019 [7]. Consequently, the World Health Organization (WHO) added vaccine hesitancy to their list of top 10 threats to global health in 2019 [31]. Vaccine hesitancy will particularly damage the containment of future outbreaks of smallpox because only a small percentage of the current populace is vaccinated, and their protection is likely to have waned.

Social media has fueled vaccine hesitancy by escalating disinformation on immunization, and in the event of an outbreak may enable malefactors who seed the disease to simultaneously amplify anti-vaccine campaigns and manipulate the target populace to refuse preventives. Also, in the age of social media, opinions regarding receptivity to vaccines rapidly evolve, through social networks that overlap with but are not identical to biological networks. Specifically, during physical interactions, both diseases and opinions may spread, whereas only opinions may spread through remote (e.g., electronic) interactions. Finally, in some cases, only disease might spread because individuals share the same physical space (e.g., public spaces like beaches, parks, public transports) without exchanging ideas. Thus, spread of smallpox and opinion regarding receptivity of vaccines must be jointly investigated.

Vaccine-hesitant individuals are likely to be more receptive towards receiving drugs. Besides, underlying medical conditions like immunocompromise contraindicate a section of the populace from receiving fast-acting live vaccines. Both vaccine-hesitant and immunodeficient (also known as immunocompromised) individuals can benefit from antiviral drugs (e.g., tecovirimat (TPOXX) and cidofovir [3]) which may be used to both treat infected persons as well as prevent disease in those exposed. These are currently stockpiled in the Strategic National Stockpile for use if there is an outbreak of smallpox in the United States [3]. Drugs have an advantage in a public health setting because they can be administered without the intervention of a healthcare professional, e.g., TPOXX is taken orally twice daily [4]. The downside though is that they need to be taken for many days throughout the outbreak. Thus, considering the comparative advantages and disadvantages of drugs vis a vis vaccines, only a combination of countermeasures may be able to successfully counter smallpox should it emerge. One can envision different policies for administering antiviral drugs: (a) Policy 1 – administer only to those who have rashes (b) Policy 2 – once the number of cases in a neighborhood exceeds a certain threshold, administer to anyone with fever or rashes in the neighborhood (c) Policy 3 – once the number of cases in a neighborhood exceeds a certain threshold, administer to everyone in the neighborhood. We therefore need a framework to assess application of a combination of countermeasure and multiple countermeasure application policies. Note that the supply of the countermeasures would be finite in practice, a constraint that ought to influence the design of smart countermeasures.

Finally, geography, spatial distribution and mobility patterns of individuals is crucial in understanding the spread of an infectious disease and the efficacy of countermeasures. Locations inhabited by the target population are comprised of different neighborhoods, some of which are adjacent, some geographically disparate. Connection between individuals in geographically close neighborhoods is expected to be more frequent than those between the disparate ones leading to heterogeneous interaction rates. Next, interactions often happen through mobility which must follow geographical constraints. That is, when individuals move from one neighborhood to another, they pass through those in-between; thus mixing between two geographically disparate neighborhoods involves mixing with individuals in the neighborhoods in between. The effectiveness of the countermeasure may well be substantially enhanced by exploiting groupings of populations in distinct locales. And, the perpetrators of the attack may exploit the geography to strategically implant the initially infected individuals, following a spatial distribution that maximizes the spread of the disease.

Mathematical modeling is an indispensable analytical tool that can help us to prepare for and respond to a smallpox incident. Several models have been constructed to understand the spread of smallpox and the efficacy of vaccination and quarantine as a response to contain the disease [14, 16, 18–20, 25, 27]. Note that immunocompetent individuals (that is, those with healthy immune systems) can be immunized by the live vaccine (ACAM2000) which provides immunity in a short time after administering if taken while they are susceptible (that is before they are infected) or in the early part of incubation (that is shortly after infection) [11, 14]. On the other hand, the immunodeficient individuals can only be given the Modified Vaccinia virus Ankara (MVA) vaccine (Imvamune) [11,14,17], MVA takes two-shots 30 days apart to provide immunity [17]. Although approximately 20% of people in the United States are immunodeficient [1,21,25], the existing work largely does not consider the impact of the immunodeficient population. But, [20, 25] considered such persons in their model, assuming though that they are contraindicated from all vaccinations and excluded them from participating in vaccination altogether. However, [11, 14, 17] have shown that immunodeficient persons can receive the MVA vaccine but it will take a longer time to provide immunity. Only a few works have considered administering drugs, e.g., [14, 19]. Evolution of opinions, mobility, impact of geography, finite supply of drugs, have not been considered even in isolation in context of evolution of smallpox. None of the existing works naturally *simultaneously* modeled these, particularly in conjunction with combination of multiple countermeasures and different application strategies.

Epidemiological investigation of other infectious diseases have considered spatial heterogeneity in the spread [6, 9]. But these have not investigated questions particularly relevant for bioterrorist attacks, namely 1)which spatial distributions of the initially infected individuals maximize the spread of the disease 2) which topological connectivities of habitations enhance vulnerability to infectious epidemics. Simultaneous administering of multiple countermeasures, multiple policies for administering them and application constraints have not been considered in models for other infectious diseases either. Last, but perhaps the most important, the joint spread of disease and opinion dynamics and the impact of one on the other remain an unchartered territory in the modeling of infectious diseases. Such opinion dynamics, encompassing receptivity to vaccine once developed and wearing protective gears like masks, is expected to strongly influence the evolution of many infectious diseases, including COVID-19, in the current age.

Our contributions are as follows. We develop a computationally tractable mathematical model that jointly captures the evolution of a smallpox incident and opinion dynamics over time and space and the impact of various combinations of spatial topologies, mobility rates, countermeasures, and strategies for their applications. The model captures the essence of stochastic evolution, while retaining computational tractability, and therefore easily scales to typical target population sizes for infectious diseases encompassing millions of individuals. Note that a Monte Carlo simulation relying on an agent based stochastic simulation will not scale to that magnitude. We utilize the model to quantify the impact of the opinion dynamics, different countermeasures and application strategies, topologies and mobility patterns on metrics that capture the overall health of the system such as total number of fatalities, and visits to health-care facilities.

Our findings lead to several policy recommendations for public health authorities: 1) We show that opinion dynamics has a substantial impact on the fatality count in the settings in which vaccines are administered. Thus public health authorities must seek to incentivize the spread of opinions favorable towards reception of vaccines over social and other media. 2) They must anticipate that bioterrorist attacks will distribute the initially infected individuals to maximize the fatalities. We identify such worst case distributions in this paper. 3) They should influence new urban designs to render them less sensitive to the spread of infectious diseases and monitor existing habitations that have specifically vulnerable topological distributions for outbreaks. We identify which habitation structures are more vulnerable to bioterrorist attacks.

4) We assess the relative efficacy of different countermeasures and conclude that a combination of vaccines and antiviral drugs is the most effective in reducing the fatalities, but if one must choose one countermeasure, in this particular case, it ought to be the drugs. Accordingly, we assess the efficacies of three separate policies for administering the drugs and identify the most effective one for various parameter combinations. We show that when the availability of the drug is finite, increase in its supply substantially reduces overall fatality. Thus public health authorities should stock adequate amounts of antiviral drugs to contain future outbreaks and choose policies for administering them in accordance with their available supply. Finally, we expect that our model would easily port to other infectious diseases such as COVID-19 through the consideration of a different set of disease states and parameters.

## Methods

### Developing the model - state transition formulations

Infectious diseases evolve in different stages, different stages show different symptoms and the initial stages need not show any symptom. In smallpox, individuals move from susceptible to early incubation to late incubation to prodrome to early then late rash. From late rash, patients either recover or die. Different stages have different durations. The incubation phase typically lasts 8 to 17 days and does not have any symptoms. The prodromal phase begins at the onset of the first symptoms (fever, chills, headache) and lasts for 3 days on average. The early and late rash periods last for 3 and 7 days respectively [14]. We use *susceptibles, incubators, prodromals* to denote the individuals in susceptible, incubation and prodrome stages respectively. Patients can infect susceptible individuals in the prodrome, early and late rash stages. Thus, note that even when patients do not show symptoms they can infect others.

Efficacy of countermeasures depends upon the stage in which they are administered. Vaccines prevent the onset of smallpox with certain probabilities if administered prior to the late incubation stage. Live vaccines (ACAM2000) provide immediate immunity during this period. Immunodeficient individuals are contraindicated for receiving the live vaccine, rather they are administered the MVA vaccine (Imvamune) [11, 14, 17]. MVA takes two-shots 30 days apart to provide immunity [17]. Antiviral drugs like tecovirimat (TPOXX) and cidofovir [3] are generally administered to infected persons for treatment and can be used for prevention.

The application of countermeasures introduces additional states, namely preempted. Individuals enter the preempted state after developing immunity owing to the reception of vaccines. Antiviral drugs preempt smallpox, so individuals also enter this state if they are administered the drug. We assume that once individuals without symptoms are administered this drug, to prevent the onset of smallpox even after possible exposure, they continue to receive the drug until the disease is completely contained. Thus, preemption is permanent, and the preemption state is an absorbing state, i.e., individuals can only enter this state, not leave it.

We now describe the state transitions, progressively considering the following scenarios: (1) No countermeasure - neither drugs nor vaccines administered, (2) Vaccines only, (3) Drugs only, (4) Both drugs and vaccines.

### No countermeasure

We first consider the case that all individuals are in the same neighborhood, that is, they interact with each other at the same rate (homogeneous mixing). Each individual is either immuno-competent or immunodeficient. Individuals in either category may be in one of the following states: susceptible, early incubation, late incubation, prodrome, early rash, late rash, recovered, dead. Fig 1 depicts the state transitions pictorially. There are two kinds of transitions: (1) *interactional* and (2) *non-interactional*. The first kind of transition occurs as a result of physical interactions in which the biological contagion spreads, that is, when two individuals are in close proximity and one is susceptible while the other is infected (that is in one of the following states prodrome, early rash, late rash), then the susceptible is infected with a certain probability and transitions to early incubation stage. The second kind of transition occurs as a result of the natural evolution of the disease in infected individuals. For instance, the disease transition from early incubation to late incubation and from late incubation to prodrome after about 7 and 5 days respectively. From the late rash state, the immunocompetent individuals either transition to the recovered or to the dead state, while immunodeficient individuals invariably die [13].

**Fig 1:**
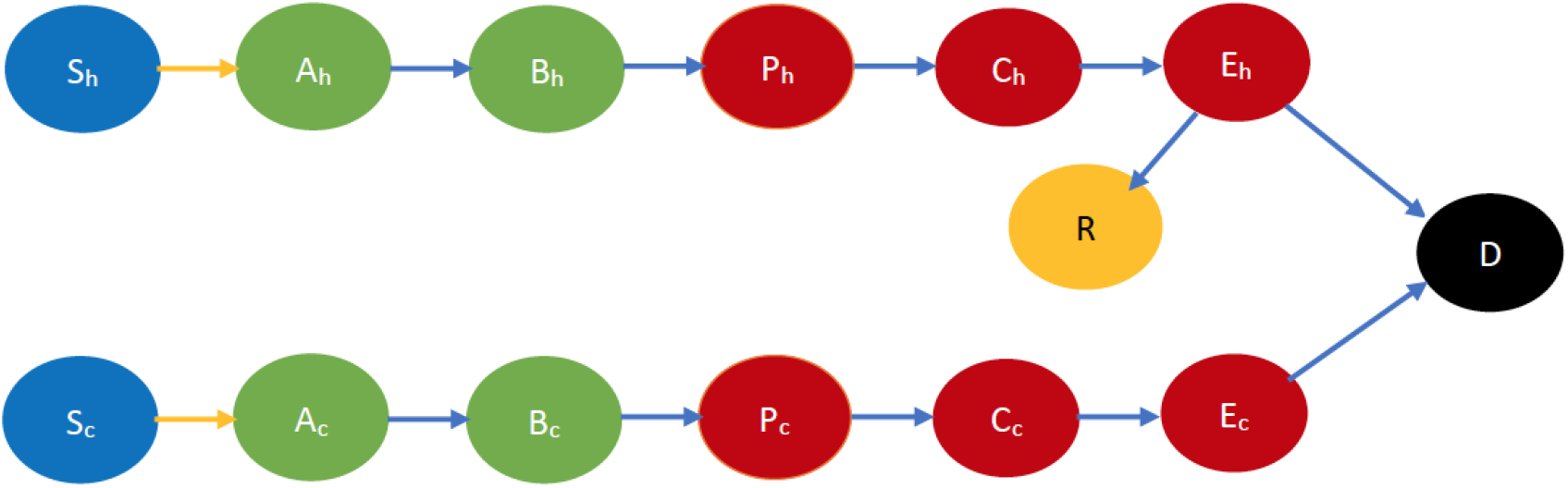
Smallpox disease progression overview when no countermeasure is implemented. The symbol S denotes susceptible, A and B denote early and late incubation respectively, P denotes Prodrome, C and E denote early and late rash respectively, R denotes recovered and D denotes dead. The suffix *h* denotes immunocompetency, and *c* denotes immunodeficiency e.g., *S*_*h*_, *S*_*c*_ respectively denote immunocompetent and immunodeficient susceptible, etc. Table 6 in Appendix A.1 contains all the relevant abbreviations we used to represent each state. The states in blue color are the susceptibles (not yet infected but they are prone to infection) while those in light green color are still in incubation period, hence they are not infectious. The states in dark red are infectious while those in gold have recovered and black denotes dead. In addition, the yellow arrows show interactional changes, namely susceptibles transitioning to the incubation state after contracting the virus from infectious individuals. The blue arrows indicate a non-interactional change, namely the natural progression of the disease.

### Drug only

The state transitions for the drug only scenario is similar to that of the no countermeasure scenario. The most important distinction here is that the disease may be preempted with the drug. More specifically, if an individual is administered the drug before he is infected he does not develop the disease while he is given the drug. If an individual is administered the drug after he is infected, he is cured of the disease with a probability that depends on the stage of the disease in which he receives the drug. We, therefore, introduce a state called *Preempted*, which we denote by *Q*, into which individuals receiving the drug transition to with the specified probability. Once an individual transition to this state, he remains in it (since he is either cured or does not develop the disease) - thus, this is an *absorbing* state. Transition to this state is non-interactional as it is induced through the consumption of drugs rather than an interaction with another individual. The state transition for the drug only scenario is depicted pictorially in Fig 2.

**Fig 2:**
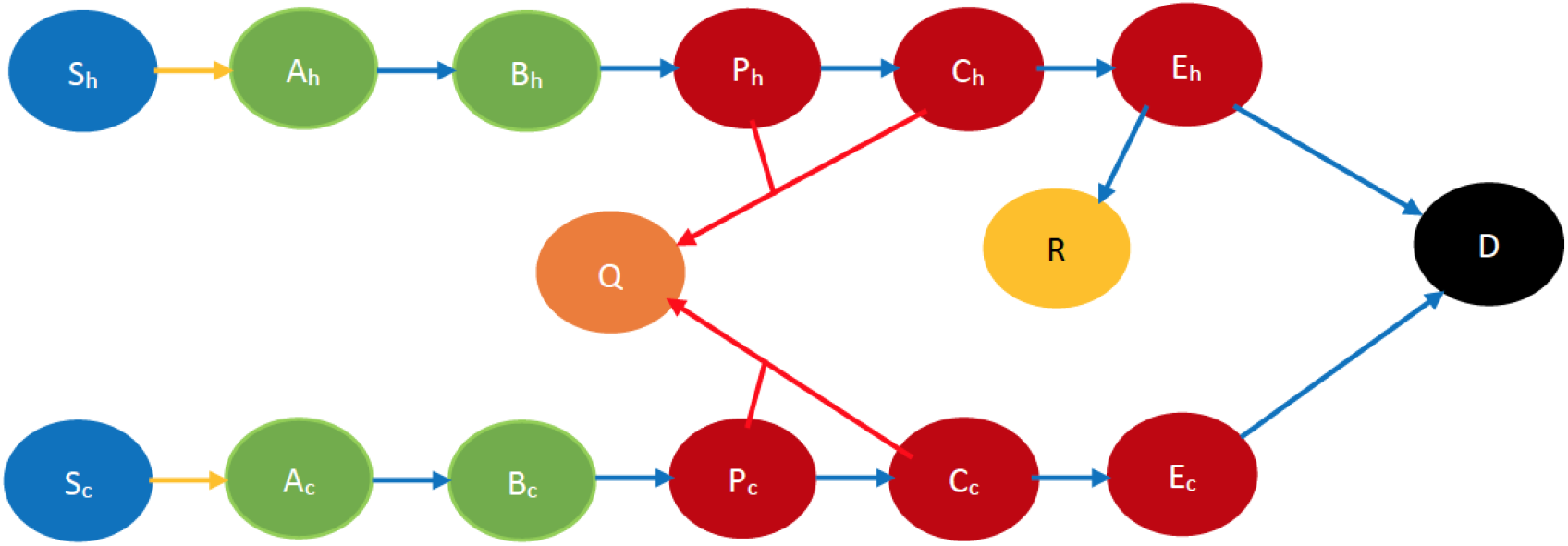
Smallpox disease progression overview for drug only scenario. The state in orange color denotes the preempted state. The red arrows denote preemption via drugs. Every other state and transition has the same meaning as described in the caption for Fig 1.

### Vaccine only

The individuals who receive vaccine transition to the preempted state, after developing permanent immunity to the infectious disease. But, some individuals may not be willing to receive vaccines. We refer to those willing to receive vaccine as *cooperative*, and the rest as *non-cooperative*. Opinion regarding cooperation evolves with interactions with other individuals. Thus, this scenario needs to model both opinion and disease dynamics and the two are coupled. Thus, the state space needs also to be enriched to consider opinions and their evolutions. Finally, one also needs to distinguish between immunocompetent and immunodeficient individuals because the latter can receive vaccines that act only very slowly regardless of their willingness to do so, and may not recover from a serious infectious disease [13].

Interactions can be of the following kinds: (a) physical interactions with an exchange of information and biological contagion (e.g., friends and acquaintances visiting homes of each other), (b) physical interactions without any exchange of ideas (e.g., people commuting on a bus, train, etc.) (c) virtual interactions with an exchange of ideas (e.g., a health worker counseling a susceptible individual over the phone or internet). (a) can cause infection and change in opinion, (b) can cause only infection, (c) can cause only change of opinion. All the above represent *interactional transitions* in this case in which both disease and opinions spread through interactions. Fig 3 depicts the state transitions of this scenario pictorially. Here we only consider the case that the cooperatives persuade the non-cooperatives to become cooperatives during opinion exchange, but in practice the opinion exchange may change opinions in the reverse direction too. In the next subsection we discuss how opinion changes in the reverse direction may be accommodated through minor modifications.

**Fig 3:**
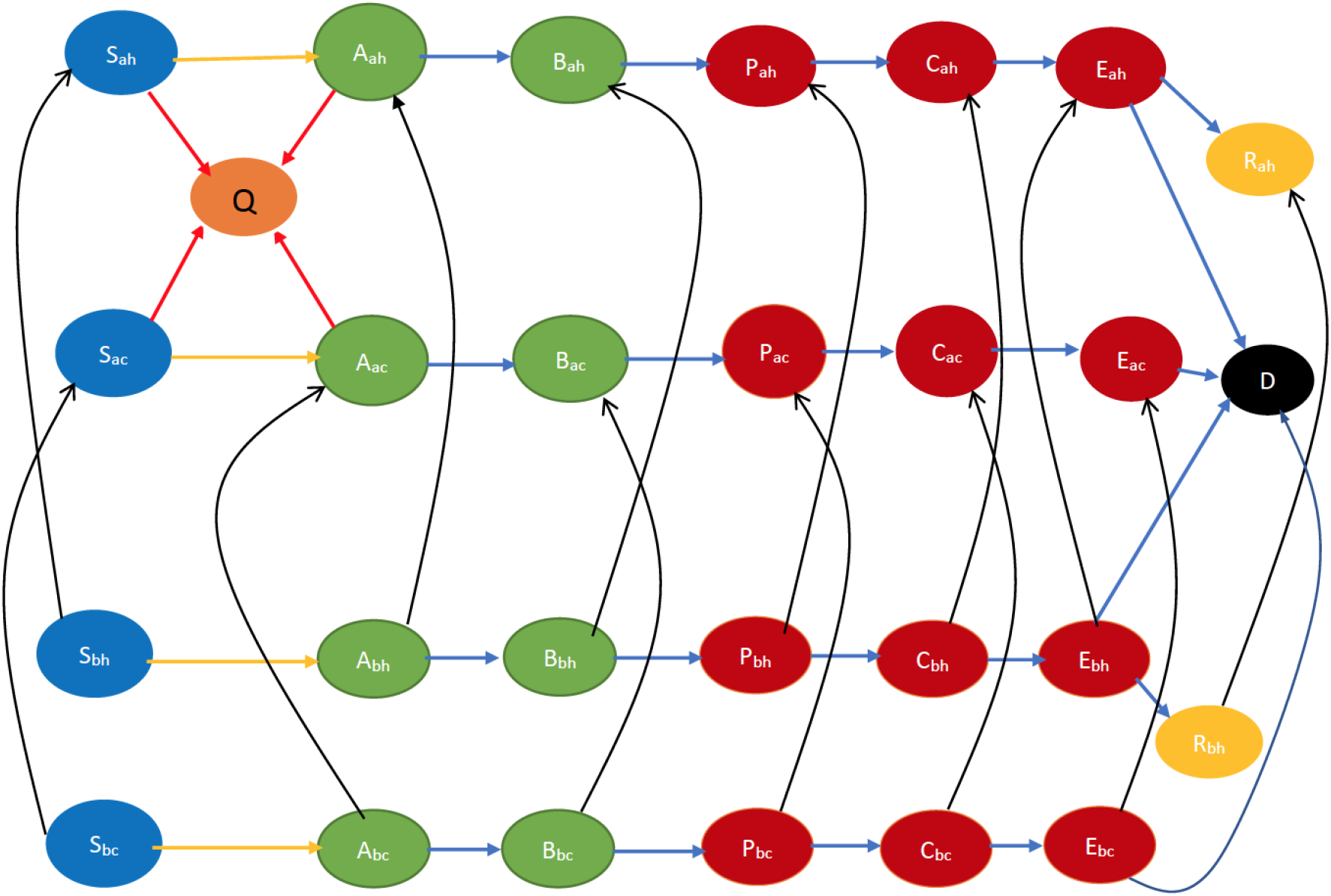
Smallpox disease progression overview for vaccine only scenario. Building on the definitions of states in Fig 1, the suffixes *a* and *b* respectively denote willingness to receive vaccine and otherwise e.g., *S*_*ah*_, *S*_*ac*_ respectively denote immunocompetent and immunodeficient susceptibles that are willing to vaccinate. Similarly, *S*_*bh*_, *S*_*bc*_ respectively denote immunocompetent and immunodeficient susceptibles that are not willing to vaccinate. Table 9 in Appendix A.3 contains all the relevant abbreviations in this scenario. The states in blue color are the susceptibles (not yet infected but they are prone to infection) while those in light green color are still in incubation period, hence they are not infectious. The states in dark red are infected and therefore infectious while those in gold have recovered, those in yellow are immunized, and black denotes dead. In addition, the yellow arrows show susceptibles transitioning to the incubation state after contracting the virus. The blue arrows indicate the natural progression of the disease. The red arrows denote preemption via vaccination while the black arrows indicate opinion evolution.

Referring to the above state transitions, we need to consider four different outcomes for interaction between individuals:

a. Neither of the individuals gets infected or changes their opinion after the interaction. For instance, let a cooperative susceptible, *S*_*ah*_, interact with a cooperative early incubator say *A*_*ah*_. This interaction does not lead to infection neither does it lead to a change in opinion about immunization.
b. Neither of the individuals gets infected, but one (and only one) changes his opinion, e.g., a cooperative susceptible, *S*_*ah*_, interacts with a non-cooperative early incubator, *A*_*bh*_. The susceptible does not contract the infection but the early incubator might change his opinion to become *A*_*ah*_.
c. One of the individuals gets infected, but neither changes his opinion. This happens for example in a physical interaction between a susceptible (e.g., *S*_*bc*_) and an infected individual (e.g., *P*_*bc*_) both of whom share the same opinion.
d. One of the individuals gets infected, and the other changes his opinion. This happens for example in a physical interaction between a susceptible (e.g., *S*_*bc*_) and an infected individual (e.g., *P*_*ah*_) who have different opinions (*S*_*bc*_ may become *S*_*ac*_ after the exchange).

### Both drugs and vaccines

An individual may be preempted by receiving either the drug or the vaccine. The preempted states are *Q*_*a*_ or *Q*_*b*_ respectively representing preempted cooperative and non-cooperative individuals. An individual can reach *Q*_*a*_ by receiving either drugs or vaccines, while he can reach *Q*_*b*_ only by receiving drug. The state transition in this case may be obtained by combining those of the drug only and vaccine only scenarios - more specifically, by adding to the vaccine-only scenario, the transitions to the *preempted state* induced by delivery of drugs. Refer to Fig 4 for the state transitions. Note that in this figure, we assume that during opinion exchange cooperatives convert non-cooperatives. To accommodate persuasion in the opposite direction, one simply needs to invert the state transition directions corresponding to opinion exchange, namely the directions of the black arrows.

**Fig 4:**
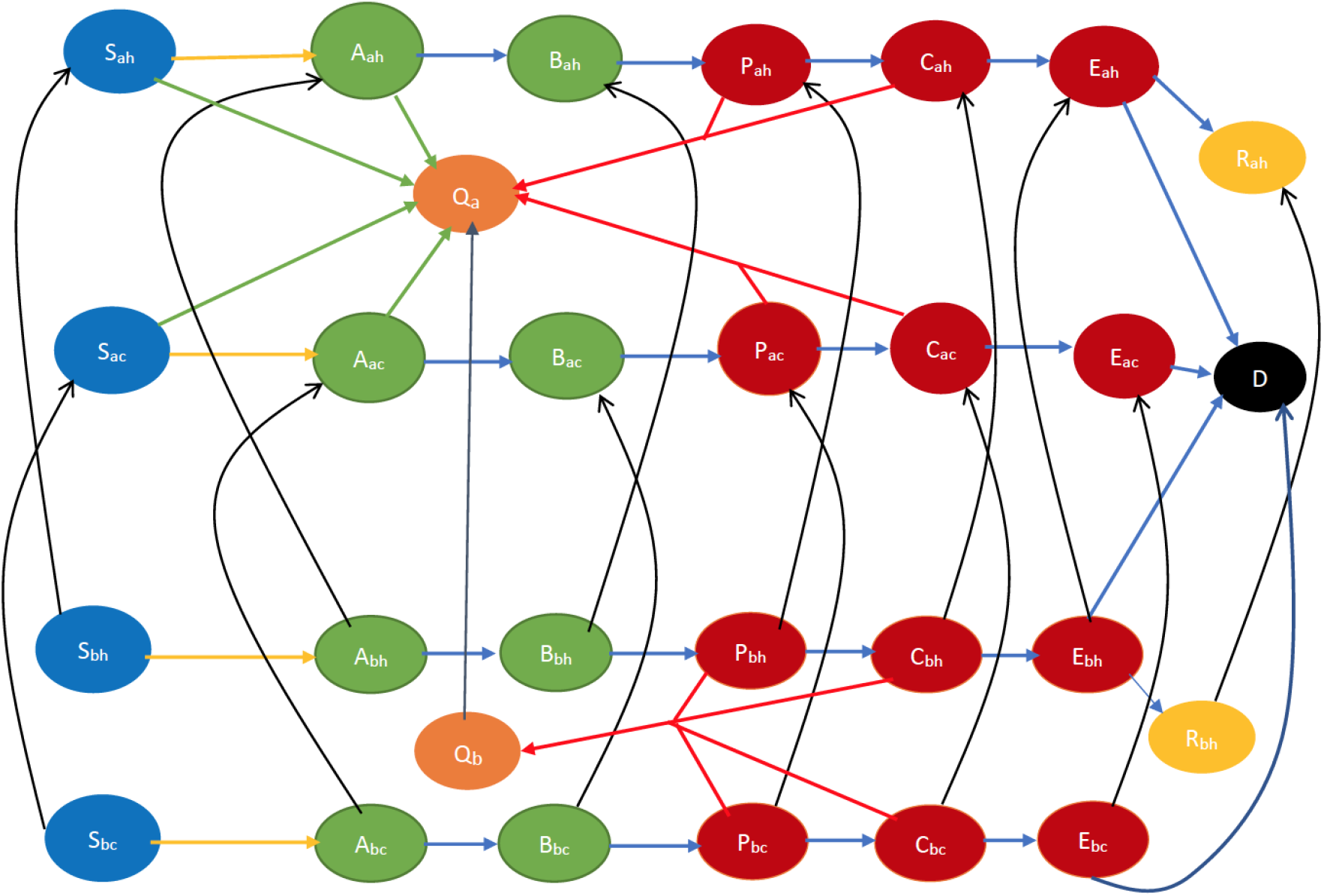
Smallpox disease progression overview for both drug and vaccine scenario. The states colored orange are the preempted states. Every other state and transition has the same meaning as described in the caption for Fig 3.

### Capturing the impact of spatial heterogeneity

We divide the target geographical area into smaller regions, referred to as *clusters*, such that the constituent regions are reasonably spatially homogeneous, that is, individuals inhabiting that region interact with each other at similar rate. The clusters can for example be neighborhoods of a city. The target geographical region can be a city, a county, a state, or even a country. People within the same cluster have high contact rates between themselves, while those living in different clusters have fewer contacts. In other words, intra-cluster interactions are higher compare to inter-cluster interactions.

So far, an individual has been characterized by his cooperativity, stage of the disease, preemption, immunocompetency or immunodeficiency. Now, he is also identified by the cluster he inhabits. Thus, the cluster becomes part of his state description, which also changes as he moves between the clusters.

To capture the geography of the region and the mobility rates of the individuals between the clusters, we represent every cluster as a node on a graph. Now, the mobility rates across the clusters may be represented through a *mobility rate matrix* on the graph. The mobility rate matrix is a matrix with rows and columns labeled by the cluster numbers, and whose *i, j* th entry represents the *mobility rate, κ*_*i,j*_, of individuals in cluster *i* to cluster *j*. Here, *κ*_*i,j*_ = 0 if it is not possible to move directly from *i* to *j* (e.g., if these are not geographically adjacent or if traffic rules do not permit vehicular mobility from *i* to *j*). If it is possible to move from *i* to *j*, then *κ*_*i,j*_ represents the probability that an individual goes to cluster *j* when he decides to move out of cluster *i* divided by the expected time he spends in cluster *i*.

We have pictorially represented some cluster decompositions in Fig 5. The central node in Fig 5a for example represents the downtown of a city, and the other nodes represent the neighborhoods surrounding it. Fig 5b depicts the cluster decomposition of a commercial and business district located at the edge of a river, sea or ocean.

**Fig 5:**
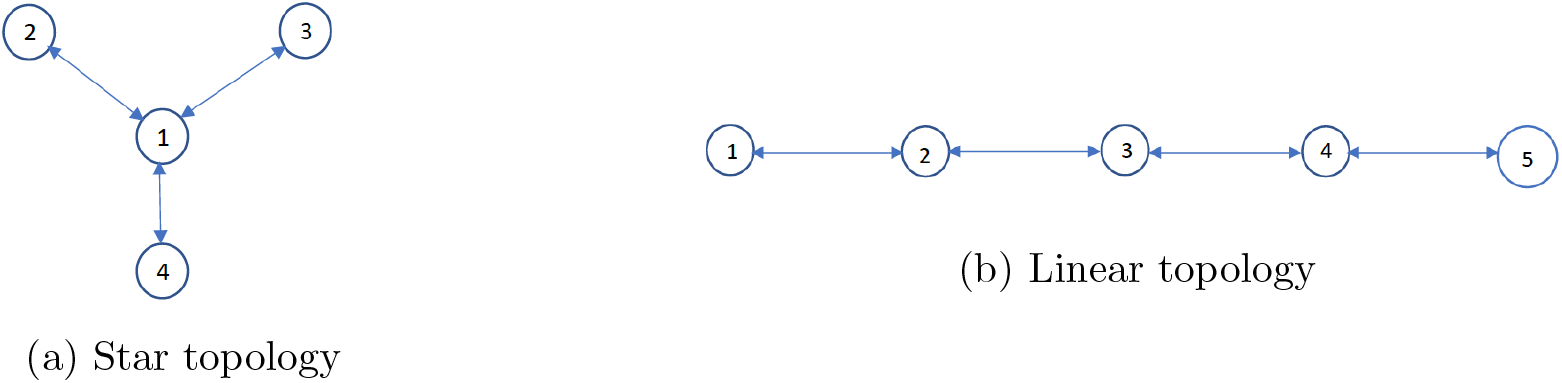
Two example topologies.

We use matrices to specify physical and virtual contact rates within and across clusters. Individuals in cluster *i* get in physical proximity of another in cluster *j* at a certain rate, the rate is much higher if *i* = *j*, than when *i* ≠ *j*. Only a fraction of these contacts spread the disease. We consider that *disease spread rate ϕ*_*i,j*_ is the product of the rate at which an individual in cluster *i* gets in physical contact with another in cluster *j* and fraction of contacts between susceptibles and infectious individuals that spread the disease to the susceptibles. Rate of an event is the expected duration between successive occurrences of the event. Typically, *ϕ*_*i,I*_ would be much higher than *ϕ*_*i,j*_ for all *i j*. Analogously, *opinion spread rate α*_*i,j*_ is the product of the rate at which an individual in cluster *i* exchanges opinions with another in cluster *j* and fraction of such exchanges that change cooperativity.

### The Clustered Epidemiological Differential Equation (CEDE) model

A joint investigation of infectious disease and opinion dynamics in presence of spatial heterogeneity inevitably leads to a computationally complex model with 1) a multiplicity of states representing a combination of stages of the disease, immunocompetency, and cooperativity, and 2) a multiplicity of state transitions representing interactional transitions due to spread of the disease and opinions, non-interactional transitions due to mobility of individuals and the natural progression of the disease in infected individuals. We model these by adapting the *metapopulation epidemiological model* [6, 9, 12, 23] which relies on a set of differential equations (co-authors of this work have utilized the metapopulation model as well [12, 23]). Metapopulation models have some distinct advantages in modeling the spread of infectious diseases. In general, estimating the spread of infectious diseases is computationally challenging because it involves millions of individuals in habitation sizes one needs to consider. Metapopulation models alleviate this challenge by relying on differential equations, which constitute computationally simple tools and can be solved using readily available numerical techniques. The differential equations capture the evolution of states of different fractions of the total population. Thus, the computational time does not increase with the increase in the size of the populace.

Metapopulation models have however thus far not captured the transitions due to spread of both disease and opinions and application of various countermeasures. We have been able to adapt the metapopulation model to capture these attributes, we describe the adaptations in Appendix A and refer to the resulting model as *clustered epidemiological differential equations* or the CEDE, given that our target area is spatially decomposed in subregions referred to as clusters.

We briefly describe the CEDE here. Each variable in the CEDE represents the fraction of the population who are in a particular system state, each state representing the combination of the cluster inhabited, the stage of the disease, immunocompetency, and cooperativity. Each differential equation captures the evolution of a particular variable. Thus, the solution of the system of differential equations provides the fraction of individuals in different states at given times, that is, the spatio-temporal distribution of the disease and opinion spread. The terms in the differential equations are either quadratic or linear. The quadratic ones represent the interactional transitions (refer to the yellow arrows in Figs 1 - 4 and the black arrows in Figs 3 and 4) and the linear ones represent the non-interactional transitions (refer to the blue arrows in Figs 1 - 4). Note that interactions always involve two individuals, hence interactional transitions are represented by quadratic terms; in contrast, the non-interactional transitions involve only one individual and are therefore represented by linear terms. This is typical of epidemiological models starting from the classical Kermack–McKendrick formulation [22]) and onward to the metapopulation models [6, 9, 12, 23]. Our work differs from the metapopulation epidemiological models in that it has additional quadratic terms representing the interactional transitions due to the spread of opinions, and additional linear terms representing preemption due to application of countermeasures.

We now provide more details on the computation time of the CEDE. Let there be *n* clusters. Then there are 29 x *n* system states in the most complex CEDE we propose, that is for the scenario in which both vaccines and drugs are administered (Appendix A.4). In this scenario, there are 29*n* differential equations and 29*n* variables. Thus, the computation time increases with the increase in the number of clusters and system states, but the rate of increase is linear in each of these. Thus, the computation time gracefully scales with the increase of the size of the system.

The CEDE is however a deterministic model, while many of the state transitions are stochastic. Nevertheless, through an application of the *strong law of large numbers*, under some commonly made regularity assumptions on the stochastic evolutions, one can show that as the number of individuals increases, the fractions of individuals in different system states in the stochastic system converge to the solutions of the CEDE, and the convergence becomes exact in the limit that the number of individuals is infinity. For the mathematical proof we refer to a recent work by one of the co-authors involving the application of the CEDE in a different domain [23]. Thus the CEDE approximates the stochastic process better as the number of individuals increase. The commonly made regulatory assumptions under which the convergence guarantees hold are that the stochastic evolutions are *Markov*, that is, the amount of time an individual spends in each system state is *exponentially* distributed, which is what we assume to estimate the parameters of the system.

The generality of the model is another important strength - it can accommodate information dynamics, arbitrary topologies, mobility patterns, countermeasure combinations, countermeasure application strategies and constraints (e.g., finite or infinite supply). Despite this generality, our model is computationally tractable and provides analytical convergence guarantees.

Finally, the model can be easily modified to accommodate diseases such as COVID-19 that spreads through contact through appropriate selection of states and parameters.

## Results

We utilized our model to obtain insights on the impact of (1) opinion dynamics as to receptivity to vaccine (2) the spatial distribution of the initially infected (3) various countermeasures (4) geography and (5) policies for administering the countermeasures on public health metrics. We mostly consider the total number of fatalities as the public health metric, but also in some cases assess the number of visits to health care facilities.

We define the fraction of individuals who are willing to vaccinate at the initial time as *initial cooperativity*. Unless otherwise stated, we use the following values of the parameters: (1) initial cooperativity is 0.80 (2) the number of individuals who are initially infected is 10, 000 (3) supply of drug is infinite (4) total population per cluster is 10 million. The rest of the parameters are chosen as in Appendix B. Unless otherwise stated, we use policy 1 for administering drugs, that is, drugs are administered to patients with rash. When we consider multiple clusters, we will consider the star and linear topology shown in Figs 5a and 5b. We consider that if clusters *i, j* are not adjacent in these topologies, the mobility rate *κ*_*i,j*_ = 0 and if they are adjacent *κ*_*i,j*_ = *κ*, where *κ* = 0.01 unless otherwise stated. For example, in Fig 5a, clusters 1, 2 are adjacent, while 2, 3 are not adjacent. Similarly, in Fig 5b, clusters 1, 2 are adjacent, while 1, 3 are not adjacent. We now consider the disease spread rate *ϕ*_*i,j*_. We assume *ϕ*_*i,j*_ = 0 if *i* ≠ *j*, that is, there is no direct physical contact and therefore no direct spread of disease between people inhabiting different clusters. We refer to *ϕ*_*i,i*_ as *ϕ*. Unless otherwise mentioned, we consider *ϕ* = 0.00173, corresponding to *R*_0_ = 6.9, the standard estimate used for smallpox. We discuss how we obtain *ϕ* from the above value of *R*_0_ in Appendix B.1. We next consider the opinion spread rate, *α*_*i,j*_. We assume that *α*_*i,j*_ = 0 if *i* ≠ *j*, that is individuals in different clusters do not directly exchange opinions. We consider that for all clusters *i, α*_*i,i*_ = *α*, and assess the system for different values of *α*, chosen from a range, rather than opting for only one estimated value. For simplicity, we will refer to *α* as the opinion spread rate henceforth. We will consider two different scenarios in our computation: 1) cooperatives convert non-cooperatives during opinion exchange (default case in our computations) and 2) non-cooperatives convert cooperatives during opinion exchange (when explicitly stated).

### Opinion dynamics has a strong impact on fatality rates

Our numerical computations in this section show that opinion dynamics have a strong impact on the fatality for the cases in which vaccines are used as a countermeasure, namely the “vaccine only” and “both vaccine and drug” scenarios. We consider two cases separately: 1) cooperatives converting non-cooperatives (Figs 6 and 7) and 2) non-cooperatives converting cooperatives (Figs 8 and 9) during interactions. In the two cases above, the cooperatives (non-cooperatives, respectively) convert the non-cooperatives (cooperatives, respectively) during their interactions at the rate *α*, thus, as *α* increases from 0 to 1, more (less, respectively) people become willing to receive vaccines, fatality in the “vaccine only” and “both vaccine and drug only” scenarios decrease (increase, respectively).

**Fig 6:**
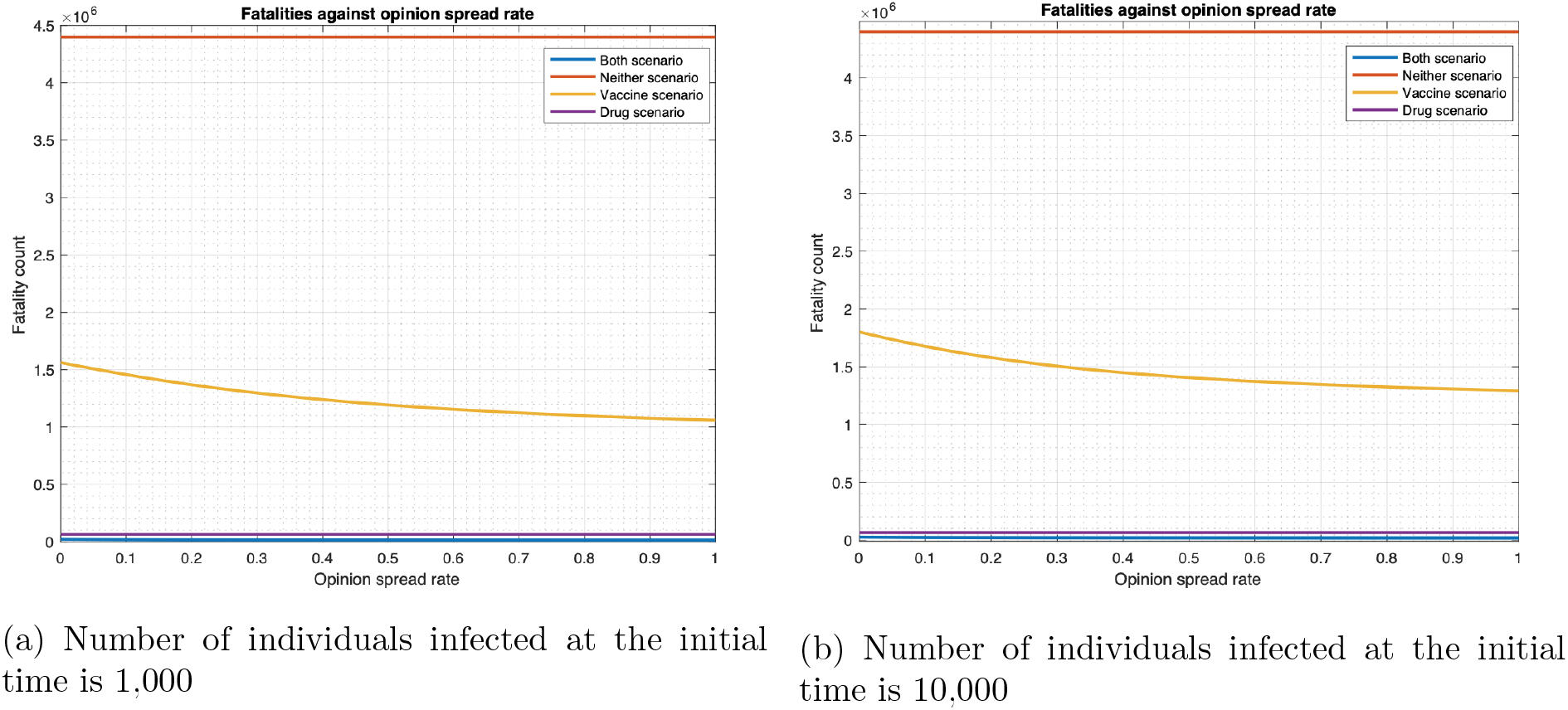
Relationship between fatality and opinion spread rate (single cluster).

**Fig 7:**
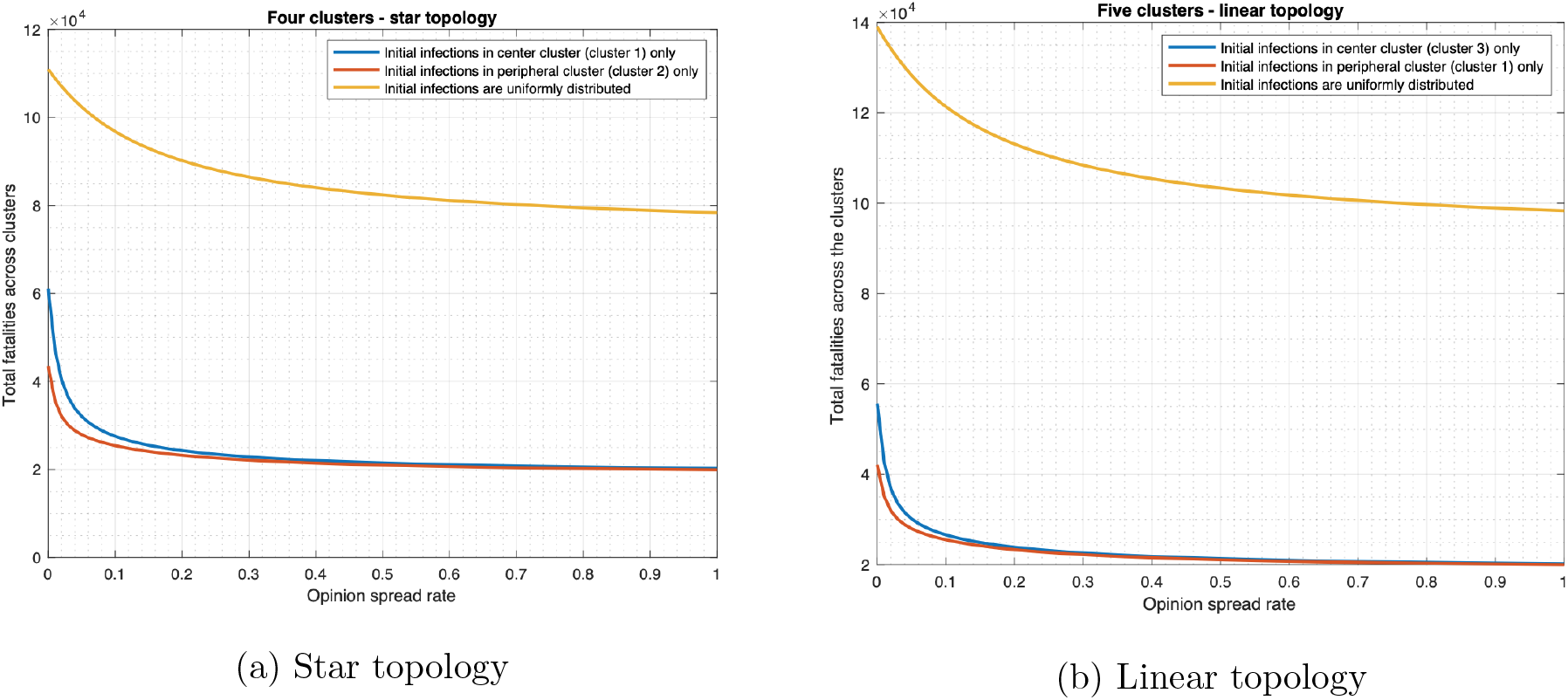
Relationship between fatality and opinion spread rate (multiple clusters).

**Fig 8:**
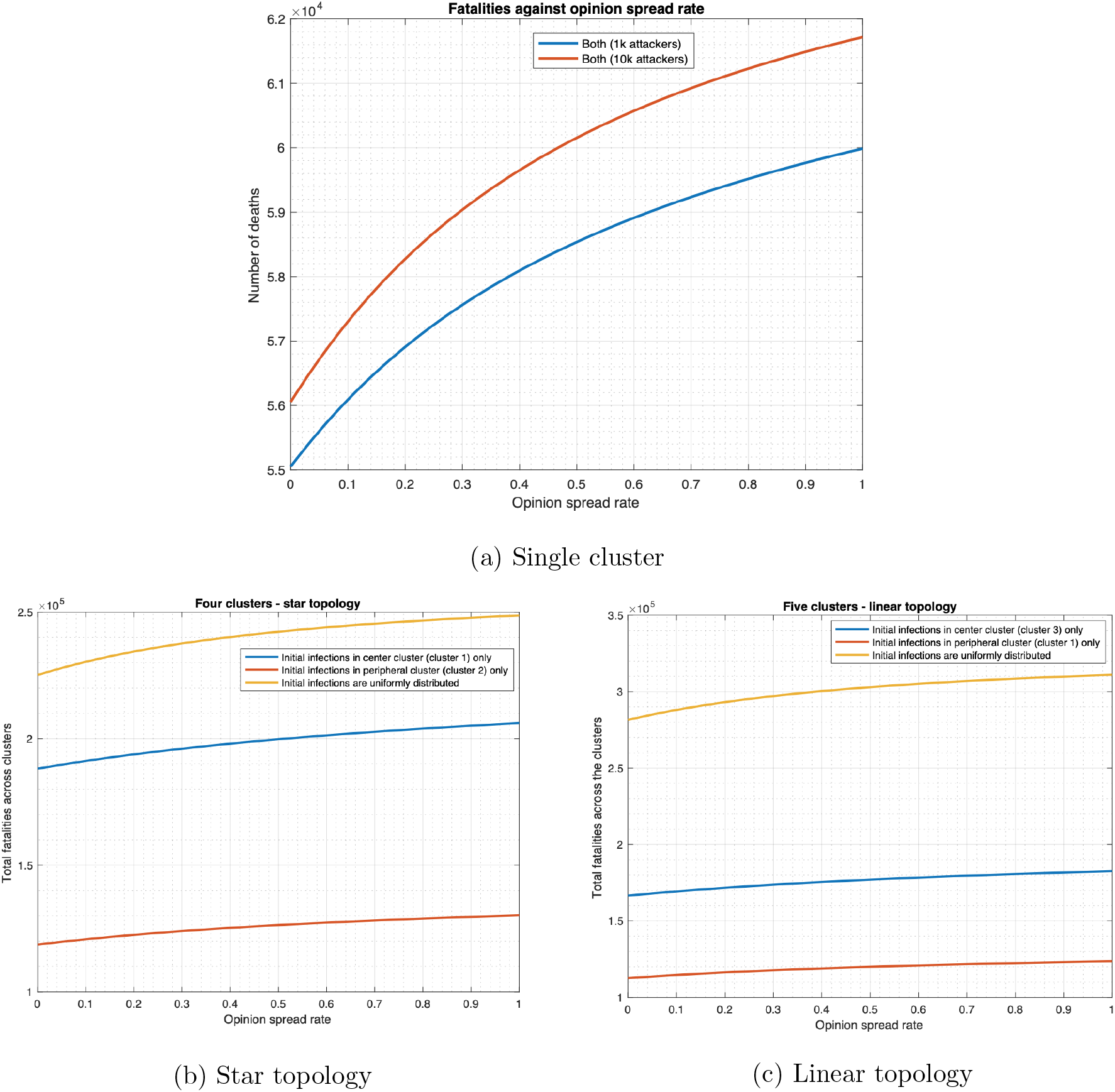
Relationship between fatality and opinion spread rate when non-cooperatives convert cooperatives.

**Fig 9:**
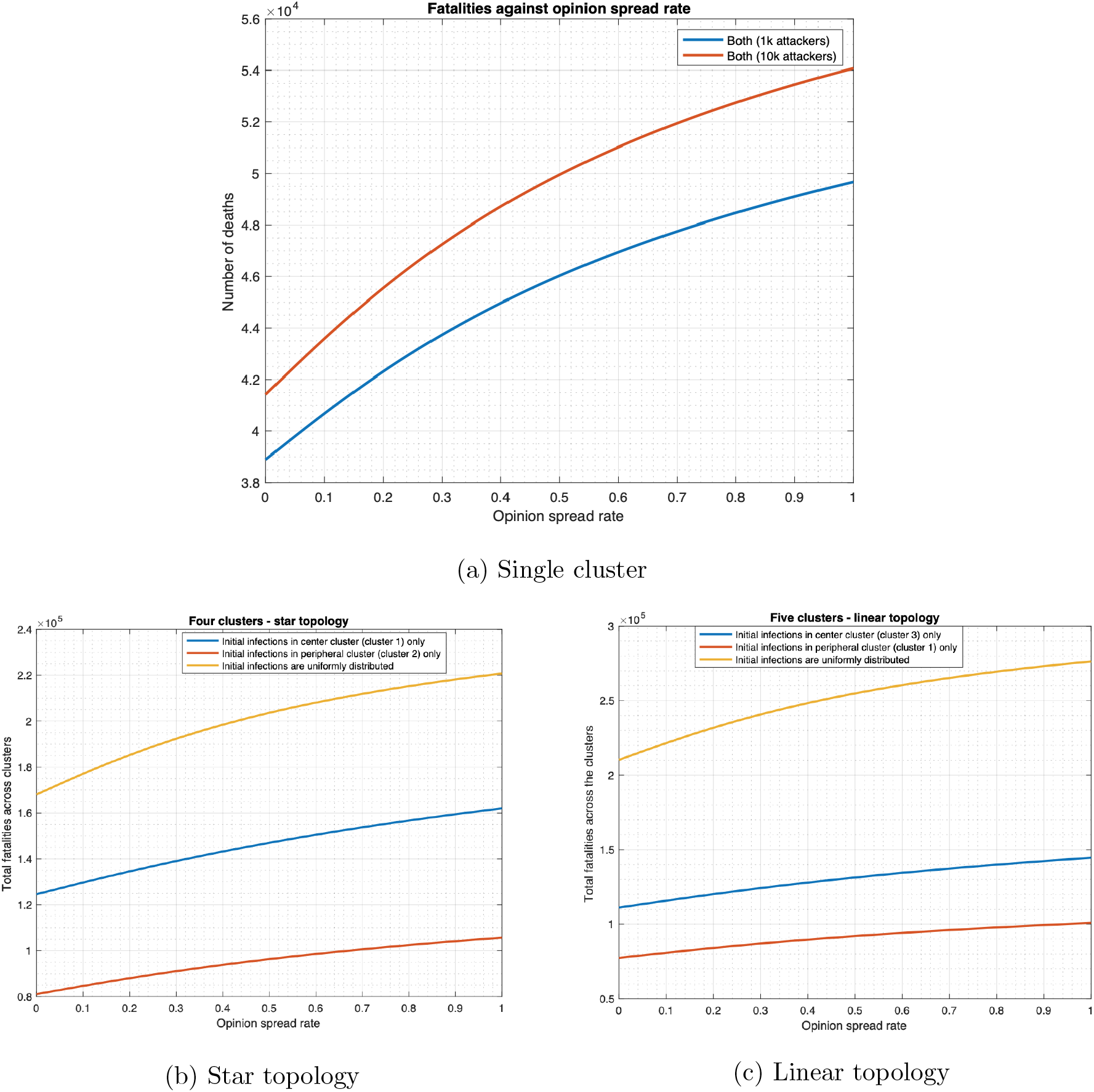
Relationship between fatality and opinion spread rate when non-cooperatives convert cooperatives (initial cooperativity = 0.5).

We start with the case that cooperatives convert non-cooperatives during interactions. Fig 6 plots the fatalities in the single cluster case in all four scenarios as a function of *α* in the range [0, 1]. With 1000 (10000, respectively) individuals initially infected, fatality for both drug and vaccine scenario decrease by 38.3% (30.7%, respectively) as *α* increases from 0 to 1. The fatality for the vaccine only scenario respectively decreases by 32.2% and 28.2% in these two cases. The fatality for no countermeasure and drug only scenarios does not change with any change in *α*.

We now consider multiple clusters for the case that cooperatives convert non-cooperatives during interactions. Fig 7 plots the total fatality for both drug and vaccine scenario as a function of *α* in the range [0, 1]. We consider three distributions of initially infected individuals across the clusters: (1) uniform distribution across clusters (2) concentration in the central cluster (clusters 1 and 3 respectively in the star and linear topologies) and (3) concentration in a peripheral cluster (clusters 2 and 1 respectively in the star and linear topologies). As *α* increases from 0 to 1 in the star topology, total fatality decreases by 29.4%, 66.8%, and 54.2% respectively in the 3 scenarios. Similarly, as *α* increases from 0 to 1 in a linear topology, total fatality decreases by 29.3%, 63.7%, and 52.4% respectively in the 3 scenarios. Therefore, the decrease in total fatality is considerable in all cases and comparable across the two topologies.

We now consider the case that non-cooperatives convert cooperatives during interactions. We report the results for the scenario in which both vaccines and drugs are administered for *α* in the range [0, 1] and for a single cluster, multiple clusters with star and linear topologies (Figs 8 and 9). As expected, with increase in *α*, fatality increases. Fig 8a shows that, in a single cluster, as *α* increases from 0 to 1, fatality increases by 8.96% and 10.10% respectively when initial number of infected individuals are 1, 000 and 10, 000 respectively. Furthermore, as *α* increases from 0 to 1 in the star topology, fatality increases by 10.45%, 9.57%, and 9.79% respectively for the 3 distributions of the initially infected enumerated in the previous paragraph (Fig 8b). Similarly, as *α* increases from 0 to 1 in the linear topology, fatality increase by 10.47%, 9.63%, and 9.83% respectively for the 3 distributions (Fig 8c). The magnitude of the change is relatively low with increase in *α* as the initial cooperativity is 0.8, so the cooperatives do not have very many non-cooperatives to convert. The magnitude of the change increases for lower values of initial cooperativity (Fig 9). For example, when the initial cooperativity is 0.5, Fig 9a shows that in a single cluster, as *α* increases from 0 to 1, fatality increases by 27.69% and 30.53% respectively when initial number of infected individuals are 1, 000 and 10, 000 respectively. Furthermore, as *α* increases from 0 to 1 in the star topology for the case where non-cooperatives convert cooperatives during interactions, fatality increase by 31.34%, 30.01%, and 30.35% respectively for the 3 distributions of the initially infected individuals (Fig 9b). Similarly, as *α* increases from 0 to 1 in the linear topology, fatality increases by 31.40%, 30.11%, and 30.40% respectively for the 3 distributions (Fig 9c).

### Distribution of initial infection and mobility of individuals have a significant impact

The outbreak of an infectious disease may start from a single cluster or multiple clusters. It is of interest to find out if fatality is higher in one of these scenarios and if the difference is significant. This would help identify how the attacks might have been designed. We also seek to understand how enhanced mobility of individuals affects fatalities.

We consider that both drugs and vaccines are used and plot the total fatality count across linear and star topologies as a function of the preemption rate. The preemption rate is the product of the reciprocal of the expected delay in developing immunity and the probability that the vaccine or drug provides immunity at the stage of the disease the individual is in. The delay is incurred in administering the vaccine or drug to the individuals (e.g., delay incurred in getting the health worker’s appointment), there is also a delay in developing immunity after the vaccine or the drug is administered. The basic preemption rates in different states have been calculated in Appendix B. We consider a preemption scale factor that multiplies all these rates, that is, scales the rates, we plot the fatality count as a function of this scale factor (Figure 10).

**Fig 10:**
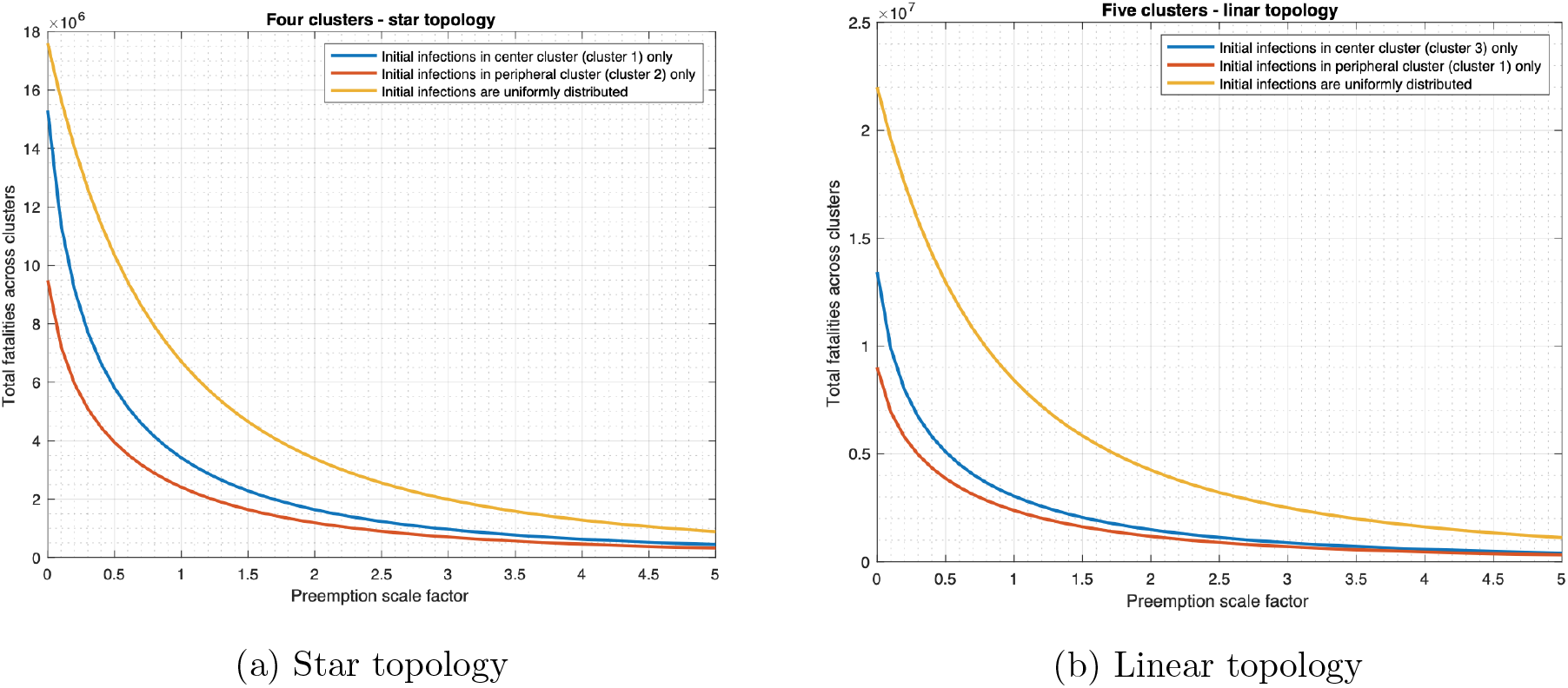
Relationship between fatality and rate of preemption.

For each topology we consider the following two scenarios:

1. Initial infections are uniformly distributed across the clusters.
2. Concentration of initial infections in a few clusters (either a central or a peripheral cluster).

Fig 10 reveals the following. Other things being equal, the fatalities are much higher when initial infections are uniformly distributed across the clusters than when all the initial infections originate from one cluster. For instance, if the preemption rate is equal to 1.6 in the star topology (see Fig 10a), then fatality when the initial infections are uniformly distributed would be 2.04 and 2.84 times the fatality when the initial infections occur only in cluster 1 and only in cluster 2 respectively. In addition, if the initial infections occur in one cluster, the fatalities are highest if the infections originate from the center. For instance, if the preemption rate in the linear topology (see Fig 10b) is equal to 0, then fatality when the initial infections originate from cluster 3 (central cluster) would be 1.49 times the fatality when the initial infections occur in cluster 1 (a peripheral cluster) only. Hence, given the same number of initial infections, the fatalities are the highest if initial infections are uniformly distributed across all the clusters and least when the initial infections originate from a peripheral cluster.

Fig 10 also shows that increasing the preemption rate substantially decreases the fatality regardless of the distribution of the initial infection. Preemption rate may be increased by decreasing the delay in administering the vaccine and delivering the drug to the individuals which may be accomplished by increasing the number of health workers and the rate of delivery of the vaccine and drugs to the health facilities.

We now plot the fatality count as a function of the mobility rates in the star and linear topologies in Figs 11a and 11b.

**Fig 11:**
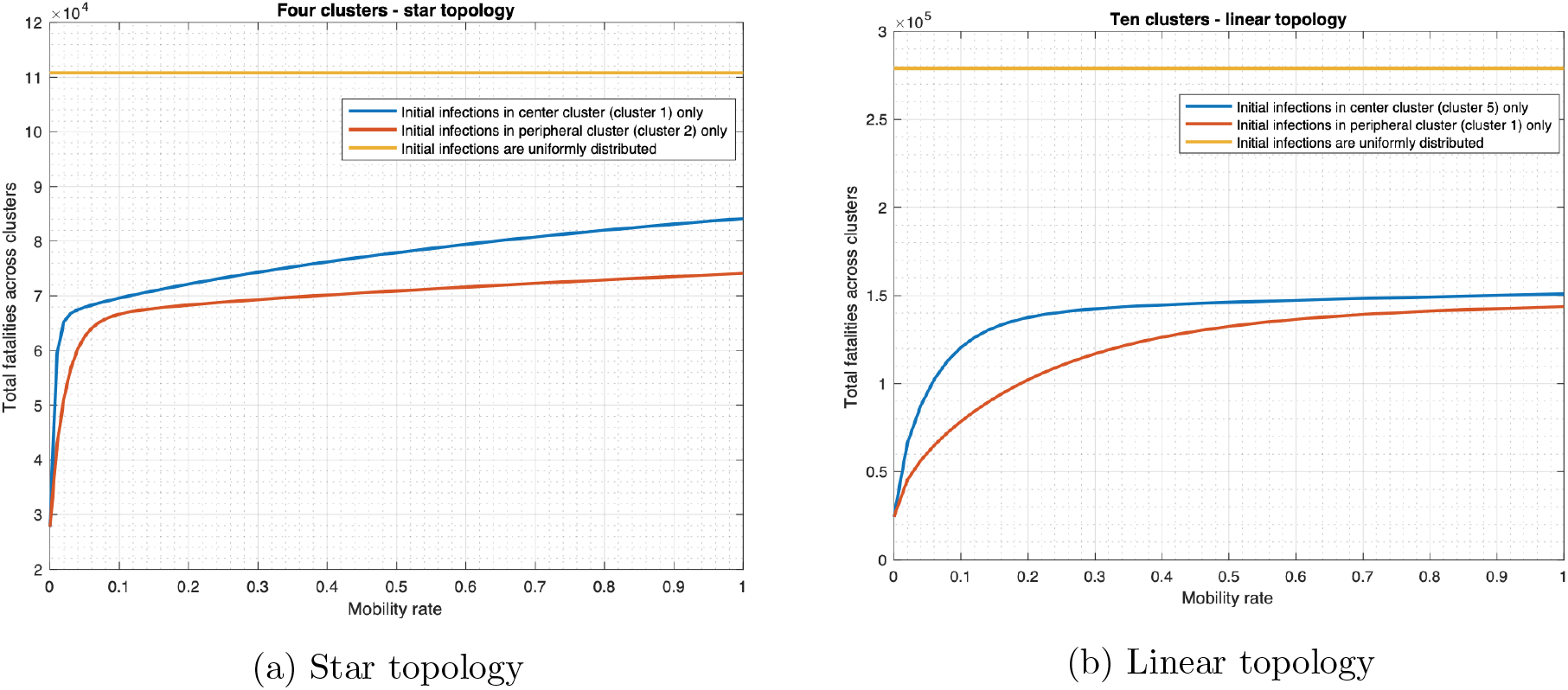
Relationship between fatality and mobility rate.

We observe the following. If the initial infections are uniformly distributed across the clusters, then fatality count does not change with the mobility rate, assuming that the mobility rates between two clusters are the same in both directions. If the initial infections occur only in one cluster, fatality substantially increases with the mobility rate. For instance, as the mobility rate increases from 0 to 1 in the star topology (Fig 11a), the total fatality increase by 202.6% (from 27, 807 to 84, 136) if the initial infections originate from the central cluster only. Similarly, the total fatality increases by 166.5% (from 27, 807 to 74, 112) when the initial infections originate from a peripheral cluster.

These observations may be explained as follows. Mobility helps individuals in different clusters mix with each other. Thus, if the initial infection is concentrated in a few clusters, higher mobility rates help spread the disease faster into other clusters and lead to high fatalities. This is the scientific basis for reducing travel links (land or air) between different regions when the infection is localized. But if the initial infection is spread out uniformly, the disease spreads in each region from the individuals initially infected even without any mobility. Thus mobility enhances the spread only marginally and thereby increases the fatality only marginally. Thus spatial quarantining strategies do not help much if the initial infection is uniformly spread. Thus, the uniform spread of initial infection constitutes the most deadly form of a deliberate attack.

We also note that regardless of the mobility rate, fatality is significantly higher when the initial infection is uniformly spread as compared to when the initial infection is concentrated in a few clusters. For instance, when the mobility rate is 0, the total fatality when initial infections are uniformly distributed across the linear topology would be 11.4 times the fatality that occurs if infections originate from one cluster (Fig 11b). When the mobility rate becomes 1, the total fatality for the uniformly distributed initial infections would be 1.9 times the fatality that occurs if initial infections occur in cluster 1 only.

### Administering drugs substantially reduces fatality

We show that using drugs as a countermeasure against smallpox substantially reduces fatality for various choices of parameter values, considering both unlimited (Figs 12 and 13) and limited (Fig 14) availability of drugs.

**Fig 12:**
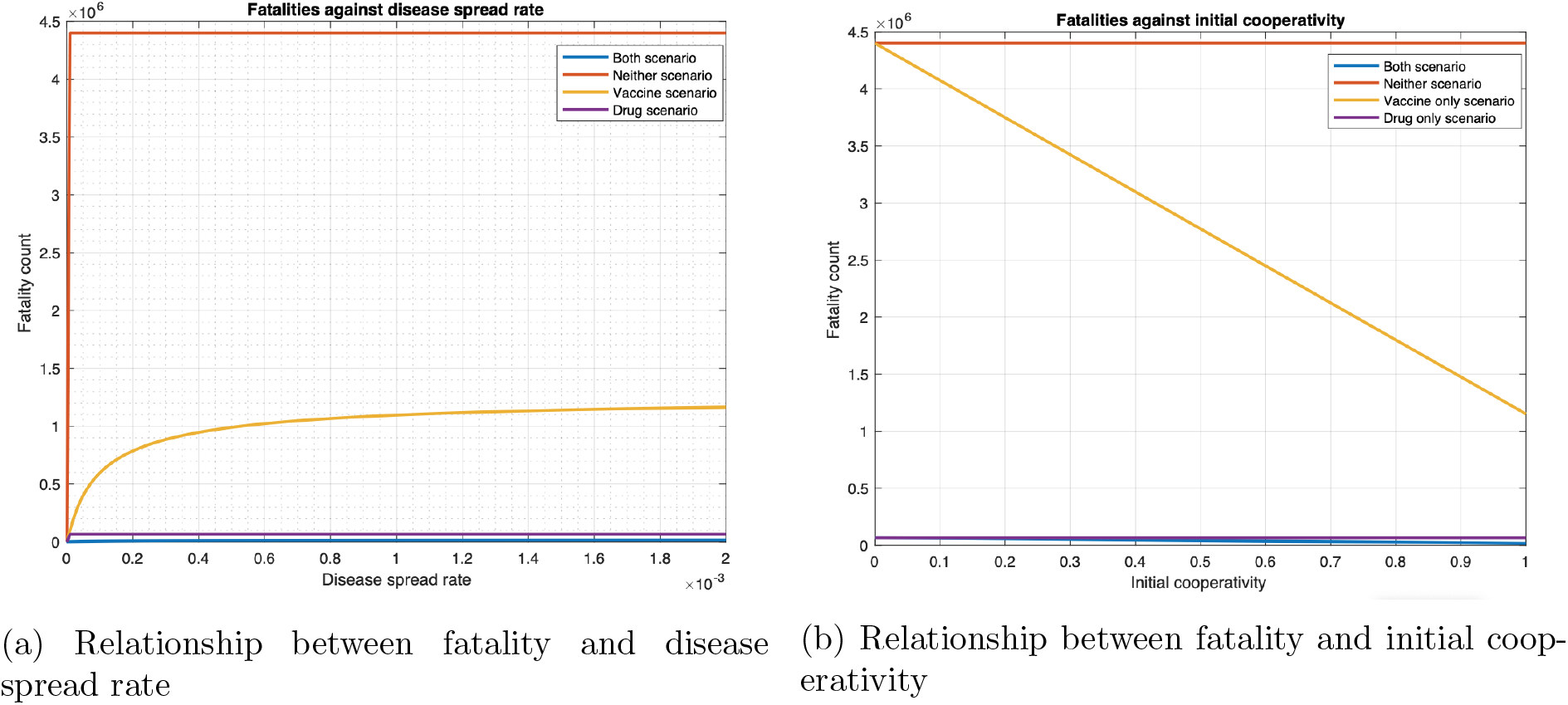
Fatalities under four countermeasure application scenarios for various values of infection spread rates and initial cooperativities for single cluster.

**Fig 13:**
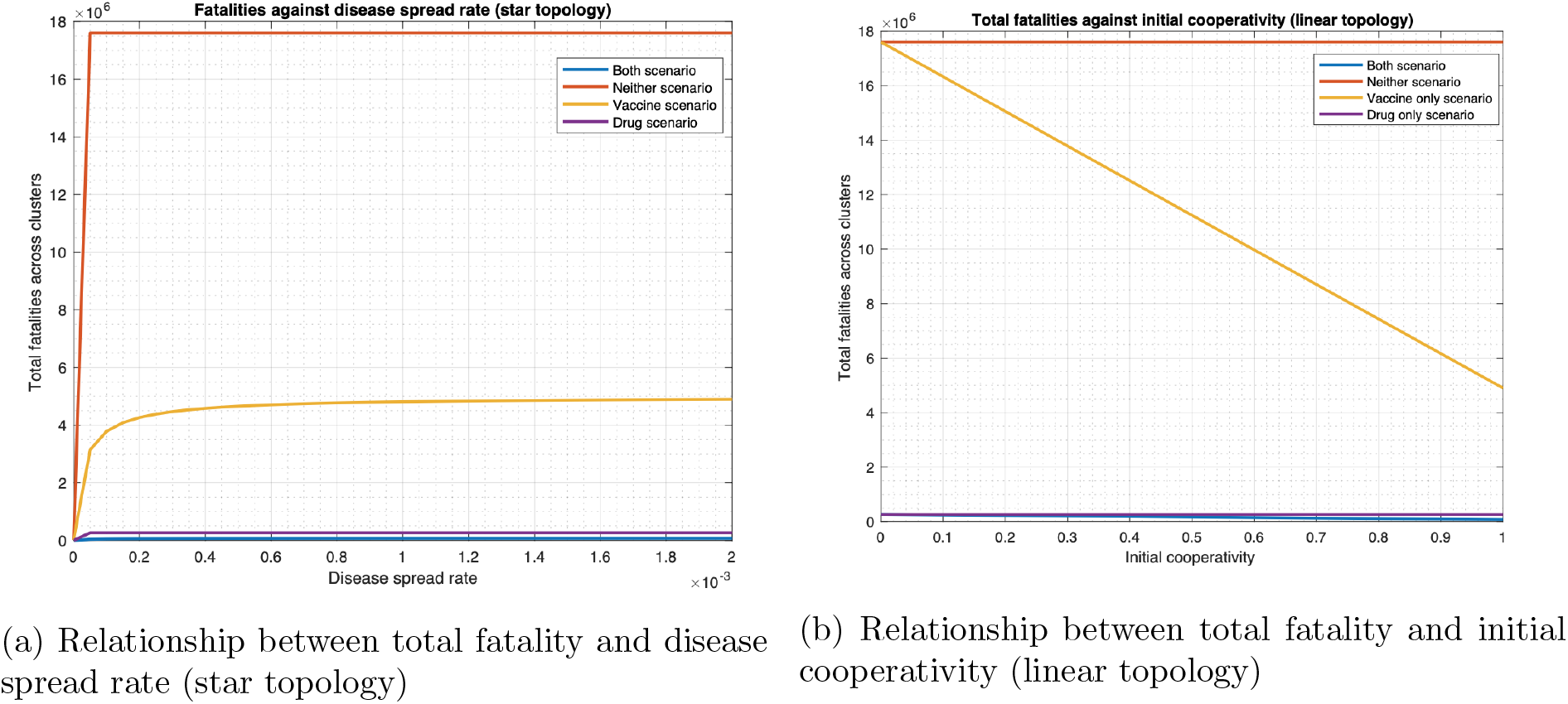
Fatalities under four countermeasure application scenarios for various values of infection spread rates and initial cooperativities for multiple clusters.

**Fig 14:**
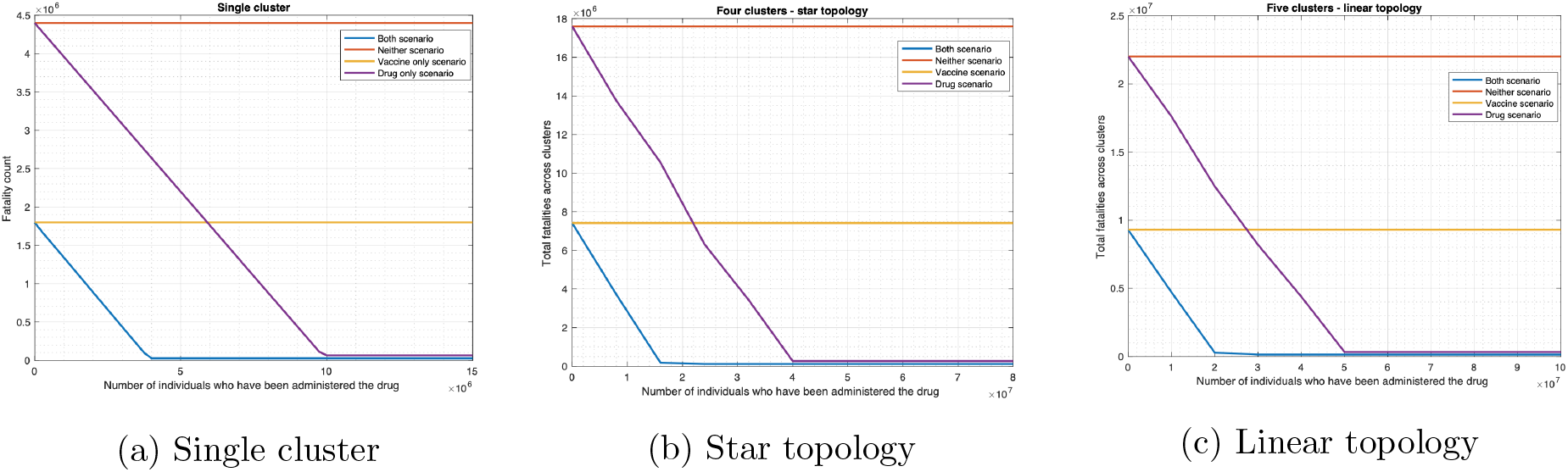
Relationship between fatalities and antiviral drug availability.

### Infinite drug availability

First, Fig 12a plots fatality as a function of disease spread rate, *ϕ* for the four scenarios for one cluster. Visually, we note that for the entire range of *ϕ* considered, the scenarios that involve administering of the drug, namely the “drug only” and “both” scenarios, have much lower fatality than those that do not involve administering of drugs, namely the “vaccine only” and “no countermeasure” scenarios. Specifically, at *ϕ* = 0.00173 (which corresponds to the standard value of *R*_0_ for smallpox as noted in Appendix B.1), the fatality for the “vaccine only” and “no countermeasure” scenarios are respectively 17.5 and 66.9 times that of the “drug only” scenario. Also, fatality for the “vaccine only” and “no countermeasure” scenarios are respectively 67.6 and 257.8 times that of the “both vaccine and drug” scenario. And fatality of the “drug only” scenario is 3.8 times that for the “both” scenario. Thus, deploying both countermeasures substantially reduce fatality, but if only one countermeasure is to be deployed, the drug is a better option in terms of fatality. This is because the non-cooperatives consent to the reception of the drug but not to the vaccine and the immunodeficient ones can only receive vaccines that act very slowly.

In addition, Fig 12b plots fatality as a function of the initial cooperativity. The conclusions with respect to efficacy of the drug remain the same as in the previous paragraph. For example, when the initial cooperativity is 0.8, then the fatality for the “vaccine only” and “no countermeasure” scenarios are respectively 27.3 and 66.9 times that of the “drug only” scenario. Also, fatality for the “vaccine only” and “no countermeasure” scenarios are respectively 67.3 and 164.5 times that of the “both vaccine and drug” scenario. And fatality of the “drug only” scenario is 2.5 times that for the “both” scenario.

We now consider the multiple cluster case. Fig 13a plots total fatality as a function of disease spread while Fig 13b plots total fatality as a function of the initial cooperativity. The patterns are similar to what we reported in the previous paragraphs. For example, when the initial cooperativity is 0.8, for the star topology, from Fig 13a, the fatality for the “vaccine only” and “no countermeasure” scenarios are respectively 18.5 and 66.86 times that of the “drug only” scenario. Also, fatality for the “vaccine only” and “no countermeasure” scenarios are respectively 66.91 and 241.28 times that of the “both vaccine and drug” scenario. And fatality of the “drug only” scenario is 3.61 times that for the “both” scenario. From Fig 13b, for linear topology, the total fatality for the “vaccine only” and “no countermeasure” scenarios are respectively 28.24 and 66.86 times that of the “drug only” scenario. Also, from Fig 13b, total fatality for the “vaccine only” and “no countermeasure” scenarios are respectively 66.95 and 158.51 times that of the “both vaccine and drug” scenario. And fatality of the “drug only” scenario is 2.37 times that for the “both” scenario.

### Finite drug availability

We now consider that the drug availability is finite and drug is no longer administered when the supply is exhausted. We assume that the first time a patient is administered the drug, he is handed out enough quantity of the drug for his daily consumption throughout the period of the outbreak. Since the period of the outbreak can vary and may depend on re-introduction of the infection from outside, we measure the drug supply in terms of the number of patients who has been administered the drug. Fig 14 plots the fatality as a function of the drug supply as measured above, and reveals that the fatality for the scenarios in which drug is administered, namely the “drug only” and “both drug and vaccine” decrease linearly with increase in drug supply, and remains constant once the supply crosses a threshold. Above the threshold value there is enough drug to administer to the population who need drugs. For the single cluster case, the fatality in the drug only case decreases from 4.4 million to 65, 808 as the drug supply increases from 0 to approximately 10 million, which constitutes the threshold value. Note that the population size is 10 million here, thus the fatality does not decrease any further once the drug supply is enough to preempt the entire population. The fatality in the both drug and vaccine scenario decreases from 1.8 million to 26, 752 (98.514%, see Fig 14a) as the drug supply increases from 0 to approximately 4 million, which constitutes the threshold value. Since individuals can also be preempted through vaccines, one only needs enough drug supply to preempt 4 million individuals to decrease the fatality count to its minimum possible value.

Similarly, for the four cluster star topology, the fatality in the drug only scenario decreases from 17.6 million to 263, 234 (98.504%, see Fig 14b) as the drug supply increases from 0 to approximately 40 million, which constitutes the threshold value. The fatality in the both drug and vaccine scenario decreases from 7.58 million to 113, 305 (98.505%, see Fig 14b) as the drug supply increases from 0 to approximately 16.4 million, which constitutes the threshold value. In addition, for the five cluster linear topology, the fatality in drug only scenario decreases from 22 million to 329, 042 (98.504%, see Fig 14c) as the drug supply increases from 0 to approximately 50 million, which constitutes the threshold value. The fatality in both drug and vaccine scenario decreases from 9.3 million to 138, 821 (98.507%, see Fig 14c) as the drug supply increases from 0 to approximately 20 million, which constitutes the threshold value. Note that for the star topology, the threshold value for “both case” is approximately 4 times that of the corresponding single cluster since there are 4 clusters. This is because the total population is 4 times the population for the corresponding single cluster. Similarly, for the linear topology, the threshold is exactly five times that for single cluster since the population size is 5 times as the number of clusters. Again, the threshold value is less than that in the drug only scenario for the same reason as for single cluster.

The fatalities for the scenarios not involving drugs naturally do not change with increase in the availability of the drug. Also, note that when the drug supply is zero, i.e., no drug is available then the fatality in the drug only scenario equals that for the no countermeasure scenario, and the fatality in both drug and vaccine scenario equals that for the vaccine only scenario. For low value of the drug supply, the vaccine only scenario has lower fatality than the drug only scenario, but as the drug supply increases, the fatality for the drug only scenario falls below that of the vaccine only scenario. Thus, if the drug supply is low and if one needs to choose between the countermeasures, vaccine is better with respect to fatality count. And, high values of the drug supply substantially reduce the fatality count.

### Impact of topology

We now compare the fatality counts across topologies and identify patterns that emerge. Towards this end, we consider the fatalities in star and linear topologies reported in Figs 7 - 14. We compare the per cluster fatality (total fatality count normalized by the number of clusters) of the star and linear topologies for representative data points in the figures and report in Tables 1 to 5. In these Tables, we compare the fatalities for three distributions of initially infected individuals across the clusters (1) uniform distribution across clusters (2) concentration in the central cluster (clusters 1 and 3 respectively in the star and linear topologies) and (3) concentration in a peripheral cluster (clusters 2 and 1 in the star and linear topologies). We represent these as “Uniform”, “Central”, “Peripheral” respectively. To distinguish between the cases that cooperatives convert non-cooperatives and non-cooperatives convert cooperatives, we add suffixes to the terms denoting the three distributions, that is, we denote the first case by UniformC, CentralC, and PeripheralC, and the second by UniformNC, CentralNC, and PeripheralNC. When we consider only the first case, which is the default, we omit the suffixes altogether. The columns titled “Normalized Linear” and “Normalized Star” respectively represent the per cluster fatalities for the Linear and Star topologies respectively. We obtain the difference between the per cluster fatalities of the star and the linear topologies (Column titled “Difference”) and present the difference as a percentage of the former in the tables (Column titled “% Change”). In Tables 1 - 4, we consider the scenario in which both drugs are vaccines are administered (“Both”). In Table 5, we additionally consider Drug only, Vaccine only, and Neither scenarios.

**Table 1:** Per cluster fatality in Figs 7 and 8. In Figs 7a and 7b, Figs 8b and 8c, we consider a star topology with 4 clusters and a linear topology with 5 clusters respectively. C in UniformC, CentralC, PeripheralC denotes cooperatives converting non-cooperatives. Similarly, NC in uniformNC, CentralNC, PeripheralNC denotes non-cooperatives converting cooperatives. Recall that *α* is the opinion spread rate. When *α* = 0, the cooperatives and non-cooperatives will have the same values.

| Distribution | Normalized Linear | Normalized Star | Difference | Percentage change |
| --- | --- | --- | --- | --- |
| Uniform/$\alpha = 0$ | 56,309 | 56,293 | -3,061 | -0.5.436% |
| UniformC/$\alpha = 1$ | 19,666 | 19,598 | -68 | -0.346% |
| UniformNC/$\alpha = 1$ | 62,208 | 62,176 | -32 | -0.051% |
| Central/$\alpha = 0$ | 33,316 | 47,038 | 13,722 | 41.19% |
| CentralC/$\alpha = 1$ | 4,041 | 5,067 | 1,026 | 25.39% |
| CentralNC/$\alpha = 1$ | 36,526 | 51,542 | 15,016 | 41.11% |
| Peripheral/$\alpha = 0$ | 22,516 | 29,657 | 7,141 | 31.72% |
| PeripheralC/$\alpha = 1$ | 4,006 | 4,978 | 972 | 24.26% |
| PeripheralNC/$\alpha = 1$ | 24,729 | 32,561 | 7,832 | 31.67% |

**Table 2:**
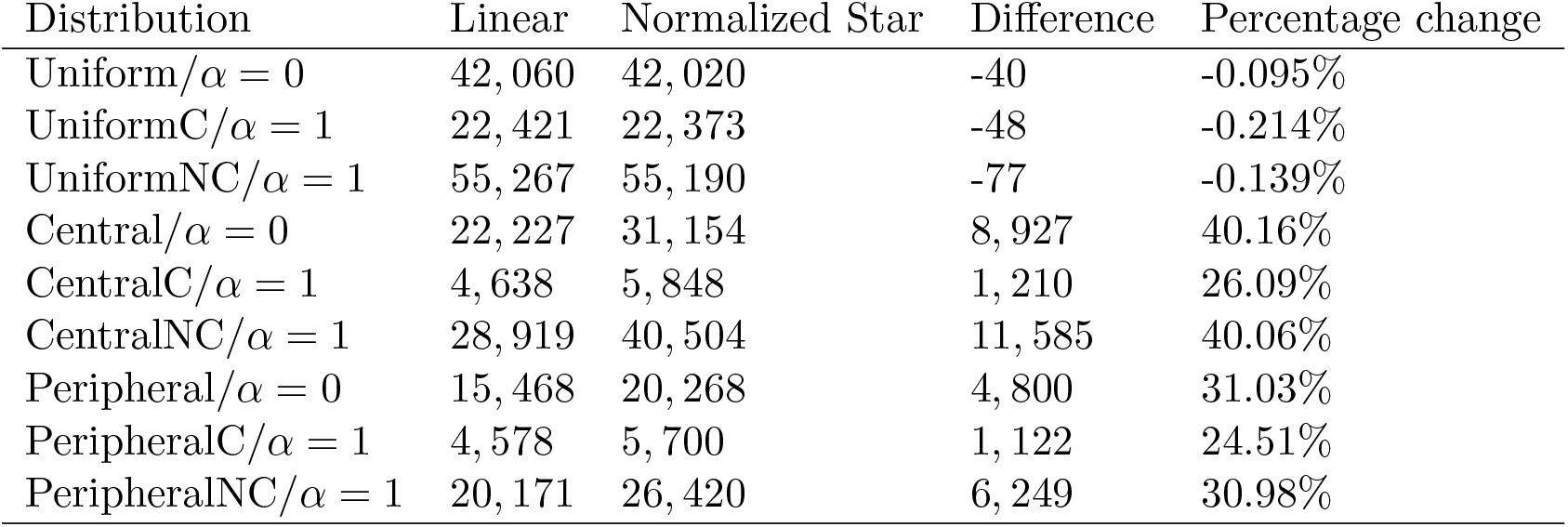
Per cluster fatality in Fig 9. We also include the values obtained for the case where cooperatives convert non-cooperatives and cooperativity is 0.5. In Fig 9b and 9c, we consider a star topology with 4 clusters and a linear topology with 5 clusters respectively. C in UniformC, CentralC, PeripheralC denotes cooperatives converting non-cooperatives. Similarly, NC in uniformNC, CentralNC, PeripheralNC denotes non-cooperatives converting cooperatives. Recall that *α* is the opinion spread rate. When *α* = 0, the cooperatives and non-cooperatives will have the same values.

**Table 3:**
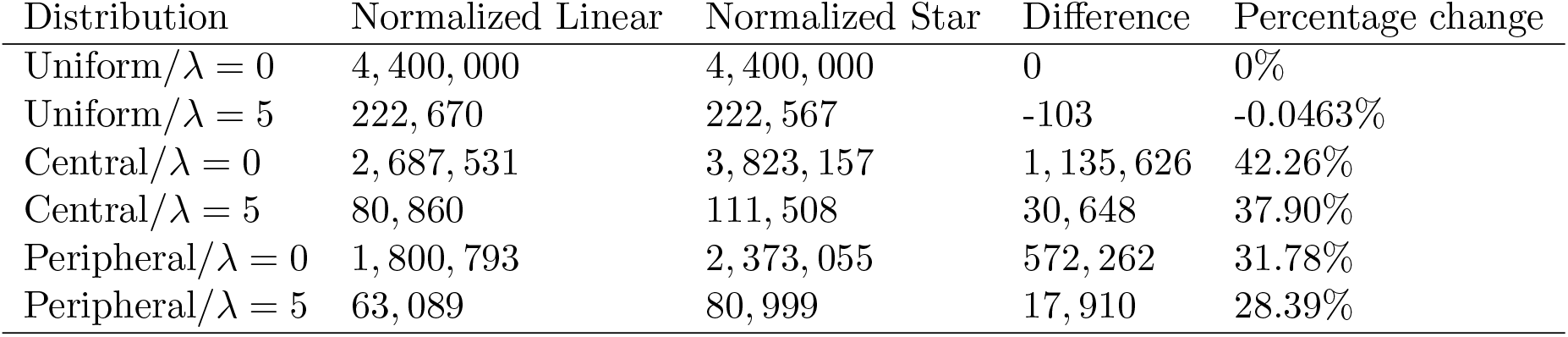
Per cluster fatality in Fig 10. In Figs 10a and 10b, we consider a star topology with 8 clusters and a linear topology with 10 clusters respectively. We denote the preemption scale factor as *λ*.

**Table 4:**
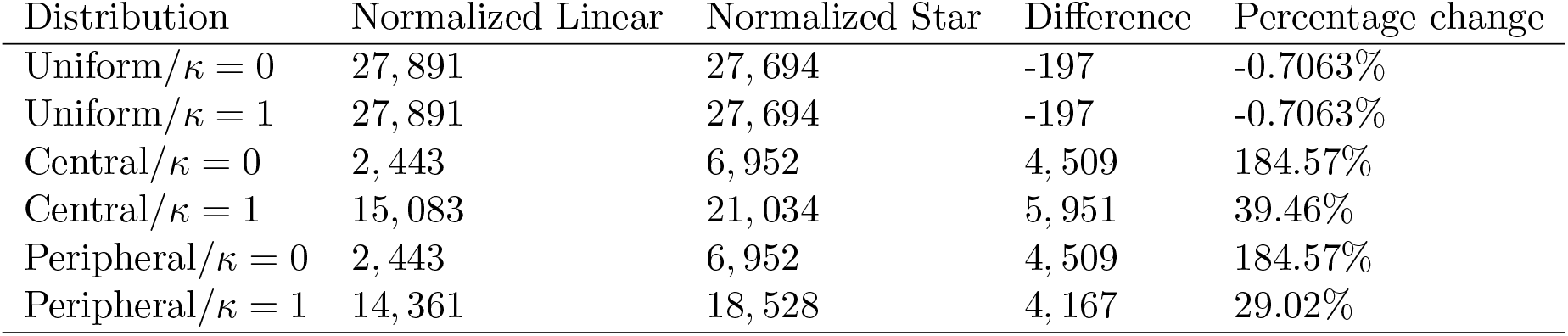
Per cluster fatality in Fig 11. In Figs 11a and 11b, we consider a star topology with 8 clusters and a linear topology with 5 clusters respectively. Recall that *κ* is the mobility rate.

**Table 5:**
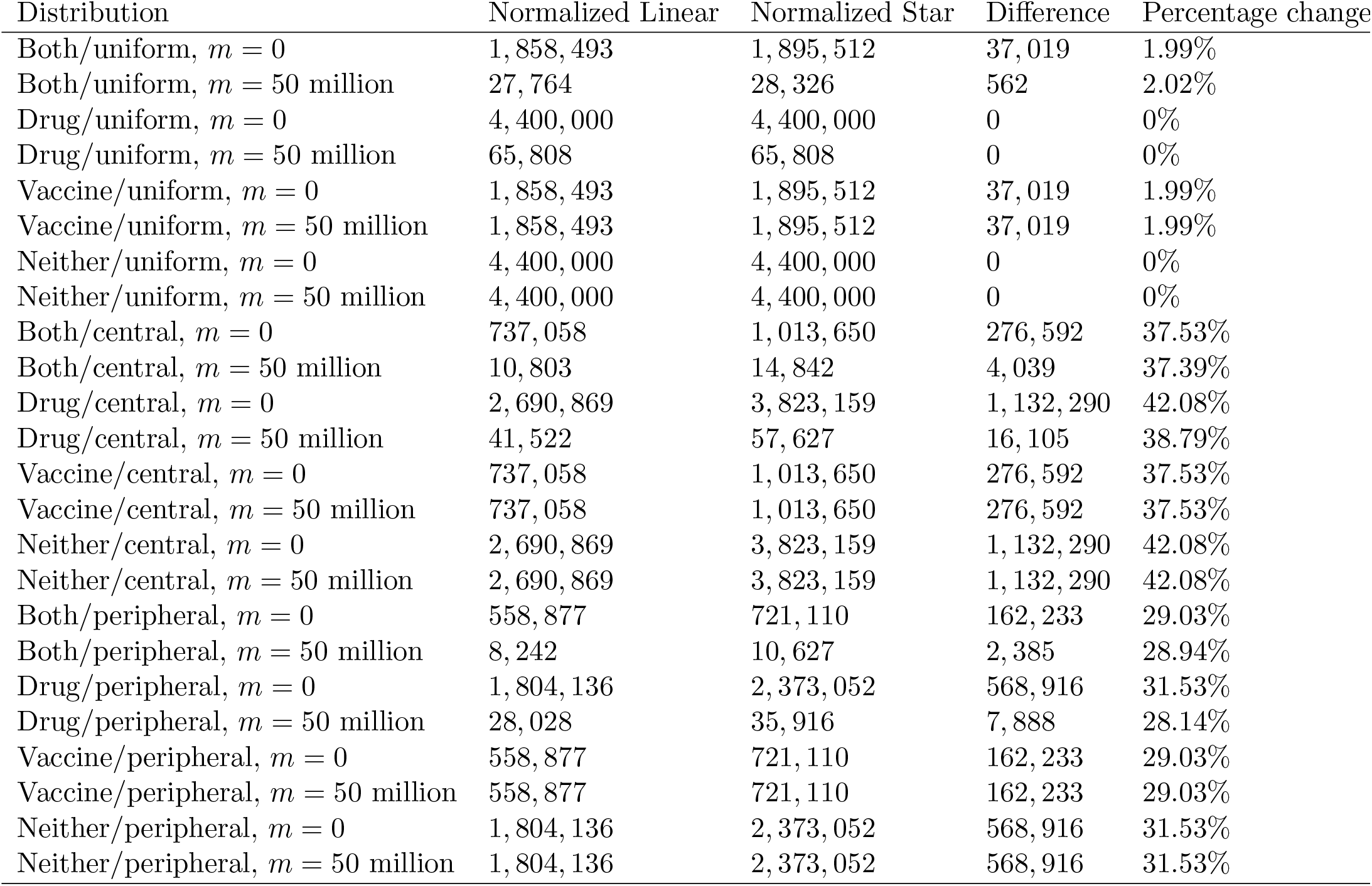
Per cluster fatality in Fig 14. In Figs 14b and 14c, we consider a star topology with 4 clusters and a linear topology with 5 clusters respectively. We denote the number of individuals who have been administered the drug as *m*.

We consistently observe the following from the tables: 1) the per cluster fatalities of the two topologies are about the same in the uniform case 2) the per cluster fatality is considerably higher in the star topology for the Central and Peripheral cases. The first happens because the initial number of infected individuals is the same in each cluster and the mobility rates to and from each cluster are the same throughout. We conjecture that the per cluster topologies will not significantly vary depending on the placement of the clusters for uniform distribution and symmetric mobility patterns. Now consider the Central and Peripheral cases. The average distance between a central or a peripheral cluster to the other clusters is higher for the star. Thus, if the initially infected individuals are concentrated in only the central or the peripheral cluster, over time the infection spreads faster in the star as compared to the linear leading to a greater number of per cluster fatalities in the former.

### Drug administering policy has a significant impact

We compare various drug administering policies with respect to (1) fatalities (2) health system load (number of doctor visits), considering the scenario in which both drugs and vaccines are administered.

We first compare policies 1 (i.e., only people with rashes receive drugs) and policy 2 (i.e., all people with fever and rash in a cluster receive drugs provided the number of patients with symptoms, that is, fever, rash, etc. exceeds a threshold of choice). First we consider only 1 cluster and threshold as 2000, that is, 0.02% of the population in each cluster. Policy 1 preempts 3, 455, 827 people with antiviral drugs while 241, 434 people died. Policy 2 preempts 3, 911, 249 people with drugs while 26, 752 people died. Thus, the fatality under policy 1 is approximately 9 times the fatality under policy 2. We get the first indication that different policies for administering drugs substantially alter the fatality count. But, clearly, policy 2 will need greater number of drugs as it preempts a greater number of individuals. We therefore next compare different policies for a finite supply of drugs.

We consider three different policies for multiple clusters and limited availability of drugs. Policies 1 and 2 are as before. Under policy 3, drugs are administered to everyone in a cluster once the number of patients with symptoms in the cluster exceeds a certain threshold (2000 for us). Drugs can be administered as long as the supply lasts. In Figs 15a and 16a, we consider the Central and Peripheral distributions for the initial infections, we consider the Uniform scenario in Figs 15b and 16b.

**Fig 15:**
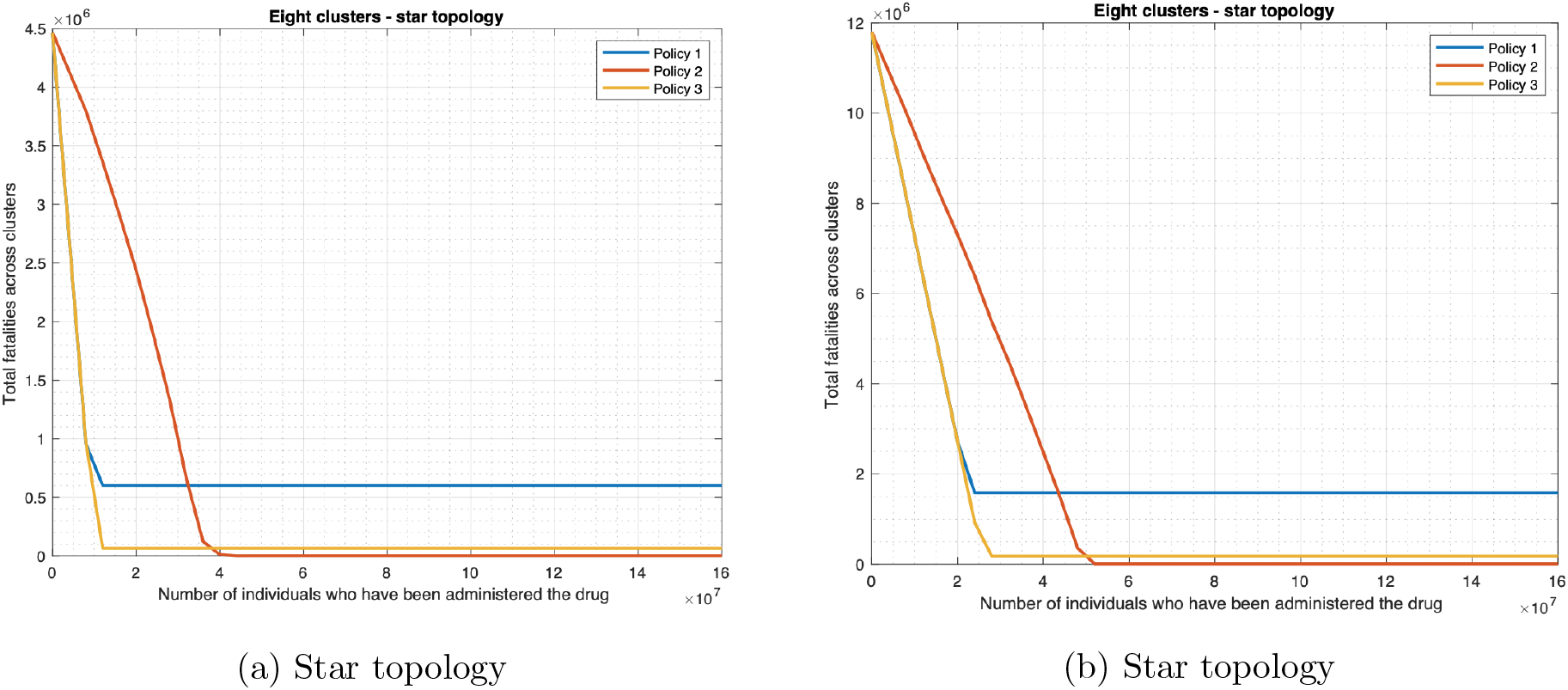
Policies for drug deployment (star topology).

We plot the fatality as a function of the drug supply as measured in the number of individuals preempted from the available supply for the star and linear topologies. We choose a star of 8 and a linear topology of 5 nodes to demonstrate the applicability of our model on topologies of different sizes. As shown in Figs 15 and 16, fatality decreases linearly with the supply of drugs until the supply reaches a threshold beyond which fatality does not change with antiviral availability. Recall that we had observed the same trend for Policy 1. We show that other policies we consider in this section follow the same trend. Next, we observe that Policy 2 always has lower fatality than policy 1. For low values of drug supply, policies 1 and 2 attain lower fatality as compared to policy 3. Thus, Policy 2 is the best in this case.

**Fig 16:**
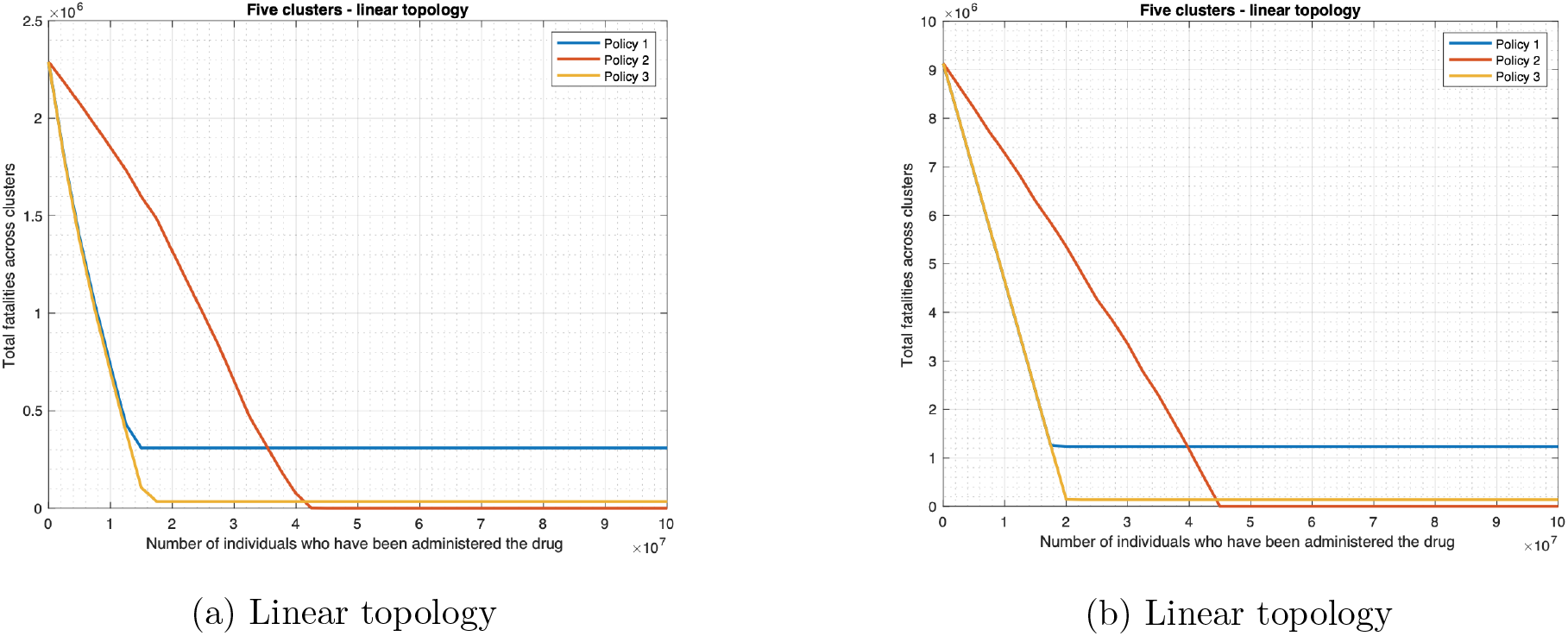
Policies for drug deployment (linear topology).

Once the drug supply exceeds a certain level, Policy 3 attains the lowest fatality among all 3 policies. Intuitively, when the availability of drugs is large, the policy that preemptively administers drugs to most individuals who are in proximity of infected individuals would attain the lowest fatalities. This is what Policy 3 does. In contrast, when the drug availability is limited, policies that administer drugs to individuals without symptoms as well will quickly exhaust their supplies and from then onwards not be able to protect those who need it most, e.g., those with symptoms. Thus, Policy 3 will not do well under these circumstances, which is what we observe. But, when the drug availability is low, it is unclear if it is better to reserve drugs for those in rash stage (Policy 1) or administer drugs to those with fever and who are in clusters where infection level has crossed a given value (Policy 2). The model informs us that the latter is a better option in this case, as by additionally preempting those with fever Policy 2 reduces the number of infectious individuals and thereby reduces the spread of the infection.

The fatalities significantly differ depending on the policy we deploy. For instance, when the initial infections are uniformly distributed across the star topology and the quantity of drugs available is enough for the population, the ratio between the fatalities of the worst and best policies at any given point can become as high as 323 (1584942 : 4902) and that between the fatalities of the second best and best policies at any given point can become as high as 36.0 (176648 : 4902) as the figures reveal.

During the outbreak of an infectious disease, the number of doctor visits of individuals is an important measure of the health of a system. We consider that an individual will visit the doctor’s office at the onset of fever (that is, Prodrome state). Thus, the number of doctor visits by individuals equals the number of individuals who enter the Prodrome state. We plot this as a function of drug supply for the three policies under consideration. As Fig 17 shows, when the drug supply is 0, the three policies will have equal number of doctor’s visits. This is anticipated as the policies differ only in the conditions under which drug is administered. The number of doctor visits decreases only slightly with an increase in drug supply for Policy 1. This is because under Policy 1 an individual is administered drug only after he develops a rash, which happens after he develops a fever, regardless of the drug supply. The slight decrease happens as greater drug supply enables the preemption of a greater number of individuals who develop rashes and thereby reduces the spread of the disease. For the other two policies, the number of doctor visits decreases considerably (and linearly) with an increase in the supply of drugs until the supply reaches a level beyond which the number of doctor visits does not change with an increase in the availability of antiviral drugs. The pattern is similar to how the fatality count changes for these policies as a function of the drug supply. And, as the drug supply increases, the number of doctor visits under Policy 1 becomes substantially higher than those for the other two policies. As shown in Fig 17a, once the drug supply exceeds a certain level, the number of doctor’s visits for Policy 1 is respectively 8.93 and 247 times that for Policy 2 and Policy 3. Similarly, from Fig 17b, the number of doctor’s visits for Policy 1 is respectively 8.97 and 323 times that for Policy 2 and Policy 3. For low values of drug supply, Policy 2 attains a lower number of doctor visits as compared to Policy 3. Thus, Policy 2 maximally reduces the load on doctors among the 3 policies in this case. Once the drug supply exceeds a certain level, Policy 3 attains the lowest load on doctors among all 3 policies. From Fig 17a, the number of doctor visits under Policy 2 is 36 times that for Policy 3 in this region. From Fig 17b, the number of doctor visits under Policy 2 is 27.7 times that for Policy 3 in this region. The explanation for this relative performance is similar to that for fatality counts given in the previous paragraph.

**Fig 17:**
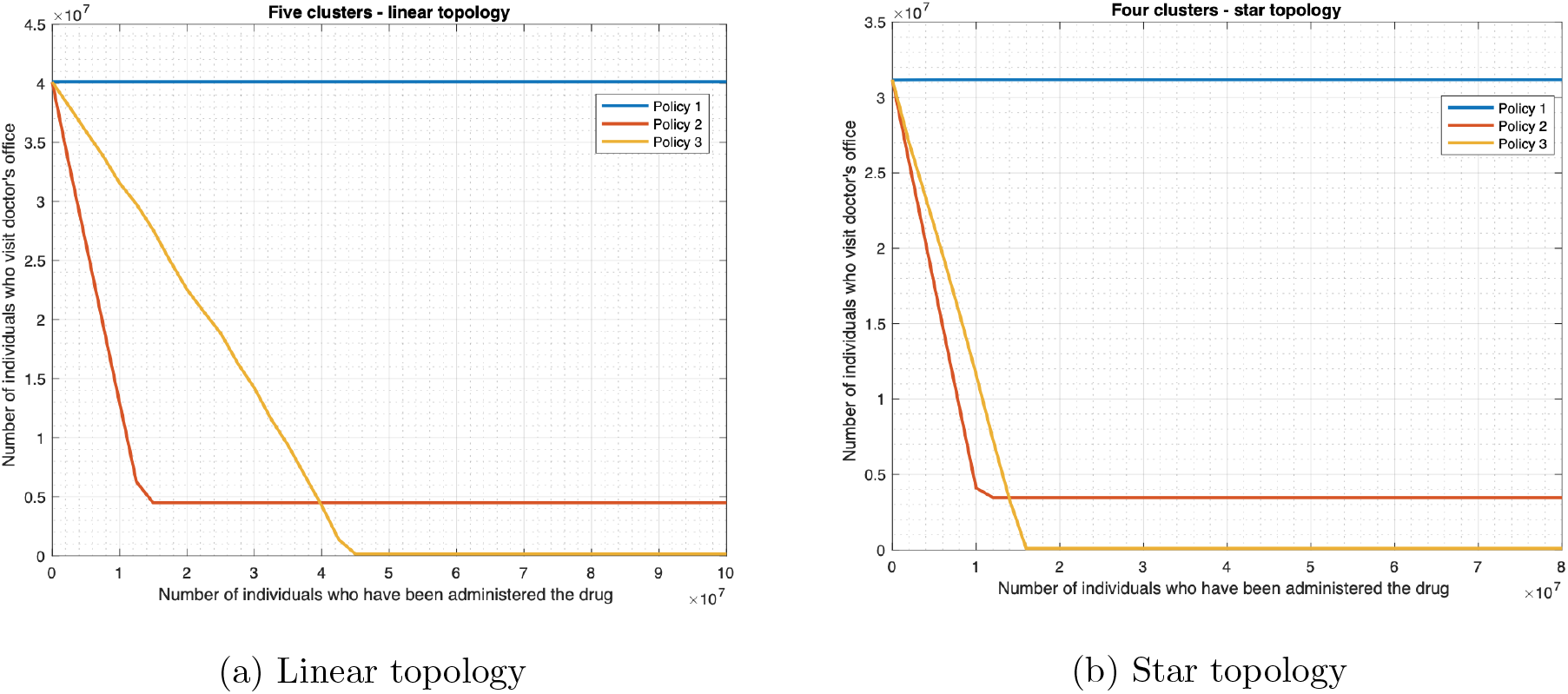
Doctor’s visits against drug availability.

## Discussions

Based on our findings, we recommend policies that public health authorities can implement should smallpox re-emerge. Some of the recommendations are specific to smallpox, others are likely to apply for the containment of any infectious disease such as COVID-19.

### From results of investigation of opinion dynamics

We have shown that evolution of opinion with respect to receptivity of vaccine has a strong impact on fatality count. Therefore, health authorities and other policymakers should seek to influence exchange of opinion towards enhancing receptivity to vaccine, possibly through health education seminars, workshops, vigorous dissemination of health information on social, digital and conventional media and through direct engagement with influencers on these platforms.

### From results of investigation on mobility patterns and distribution of initial infection

We have shown that other things being equal, epidemics are more disastrous, when 1) the initial infections are uniformly distributed across various clusters than when all the initial infections only occur in one cluster, 2) the outbreak originates from the central cluster(s) than when it originates from the peripheral cluster(s). Thus health authorities should anticipate that bioterrorist attacks will seed the initially infected uniformly across a region, the authorities should also be vigilant more about central regions than peripheral ones.

In addition, when the initial infections are uniformly distributed, fatalities in each cluster are the same irrespective of the mobility rates between the clusters. On the other hand, when the initial infections occur in one cluster only, total fatalities would escalate with an increase in mobility rates. Therefore, policymakers should opt for cordoning off areas where infection level is high before the infection spreads widely; once the latter happens reducing mobility rates across areas is unlikely to contain the disease.

### From results of investigation on impact of administering drugs

We have shown that the most effective countermeasure consists of a combination of antiviral drugs and vaccines, but if only one countermeasure can be administered it ought to be the former. Therefore, policymakers should endeavor to stock enough antivirals in case smallpox reemerges. People are usually more receptive to receiving drugs than vaccines, thus a less intense information campaign may suffice for containing the disease through the former.

In addition, we have shown that regardless of the pattern of distribution of initially infected individuals the fatalities substantially decrease if the delay in administering vaccines and delivering drugs to individuals decrease. Thus, health authorities must be prepared at short notice to supply large amounts of vaccines and drugs to healthcare facilities and deploy a large number of health workers to administer/deliver those in the event of an outbreak.

### From results of investigation on impact of topology

Our results indicate that other things being equal, the star topology has about the same or higher fatalities per cluster than the linear topology, the fatalities per cluster are about the same when the initial infections are uniformly distributed. This happens because there exists a central cluster in the star topology which is adjacent to all other topologies, in the linear topology the central cluster is much farther off from the peripheral clusters. This has important implications on pandemic sensitive urban design - topologies that have central regions that are close to all other regions will be more vulnerable to an outbreak and hence better avoided. If such topologies are inevitable because of legacy issues or because of fundamental constraints such as geography, then the health authorities need to monitor those more closely and stock greater amount of drugs for the residents and incentivize formation of consensus to accept vaccines as and when available.

### From results of investigation on various drug administering policies

We find that for low (large, respectively) values of drug supply, the health authorities should opt for policy 2 (3, respectively) to lower the fatality count and the number of visits to doctors’ offices. Thus, policymakers should be flexible about the choice of the policy and adapt it to the resource at hand.

*Our first, second and fourth findings and the resulting recommendations are also likely to hold for other infectious diseases, e.g*., *COVID-19. The other two findings and the resulting recommendations are specific to smallpox*.

## Data Availability

Detailed explanation of how all data referred to in the manuscript were estimated are provided in the appendix with full references.

## Acknowledgment

SIGA Technologies, Inc. has provided gift to support the Sarkar lab.

## A Equations of Smallpox disease dynamics

### A.1 Developing the Clustered Epidemiological Differential Equations for Disease, Information and Countermeasure Dynamics

We first consider the case of no countermeasure.

Referring to the notations in Table 6, we index each term by the clusters the individuals in the state inhabit. More specifically, *S*_*hi*_(*t*) is the fraction of individuals who are susceptible and immunocompetent and are in cluster *i* at time *t, S*_*ci*_(*t*), *A*_*hi*_(*t*), *A*_*ci*_(*t*), *B*_*hi*_(*t*), *B*_*ci*_(*t*), *P*_*hi*_(*t*), *P*_*ci*_(*t*), *C*_*hi*_(*t*), *C*_*ci*_(*t*) may be defined similarly using Table 6 as a basis. Finally, *R*_*i*_(*t*), *D*_*i*_(*t*) are respectively the fractions of individuals who are recovered and dead and are in cluster *i* at time *t*. Let *I*_*i*_(*t*) be the fraction of the *infectious* individuals in cluster *i* at time *t*, that is, the individuals who can pass on the disease to the susceptibles through contact. Then,

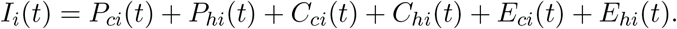

Interaction between individuals corresponding to *S*_*hi*_(*t*), *S*_*ci*_(*t*) and *I*_*j*_(*t*) spread the disease to the susceptibles and transform them to early incubators - refer to the yellow arrows in Fig 1. Natural progression of the disease change *A*_*hi*_(*t*) to *B*_*hi*_(*t*) to *P*_*hi*_(*t*) to *C*_*hi*_(*t*) to *E*_*hi*_(*t*), and finally *E*_*hi*_(*t*) to either *R*_*i*_(*t*) or *D*_*i*_(*t*); also, *A*_*ci*_(*t*) to *B*_*ci*_(*t*) to *P*_*ci*_(*t*) to *C*_*ci*_(*t*) to *E*_*ci*_(*t*), and finally *E*_*ci*_(*t*) to either *R*_*i*_(*t*) or *D*_*i*_(*t*) - refer to the blue arrows in Fig 1. Mobility changes the cluster indices. The transitions are similar for the immunocompetent and immunodeficient individuals, but the fractions of those who recover in each populace are different.

**Table 6:**
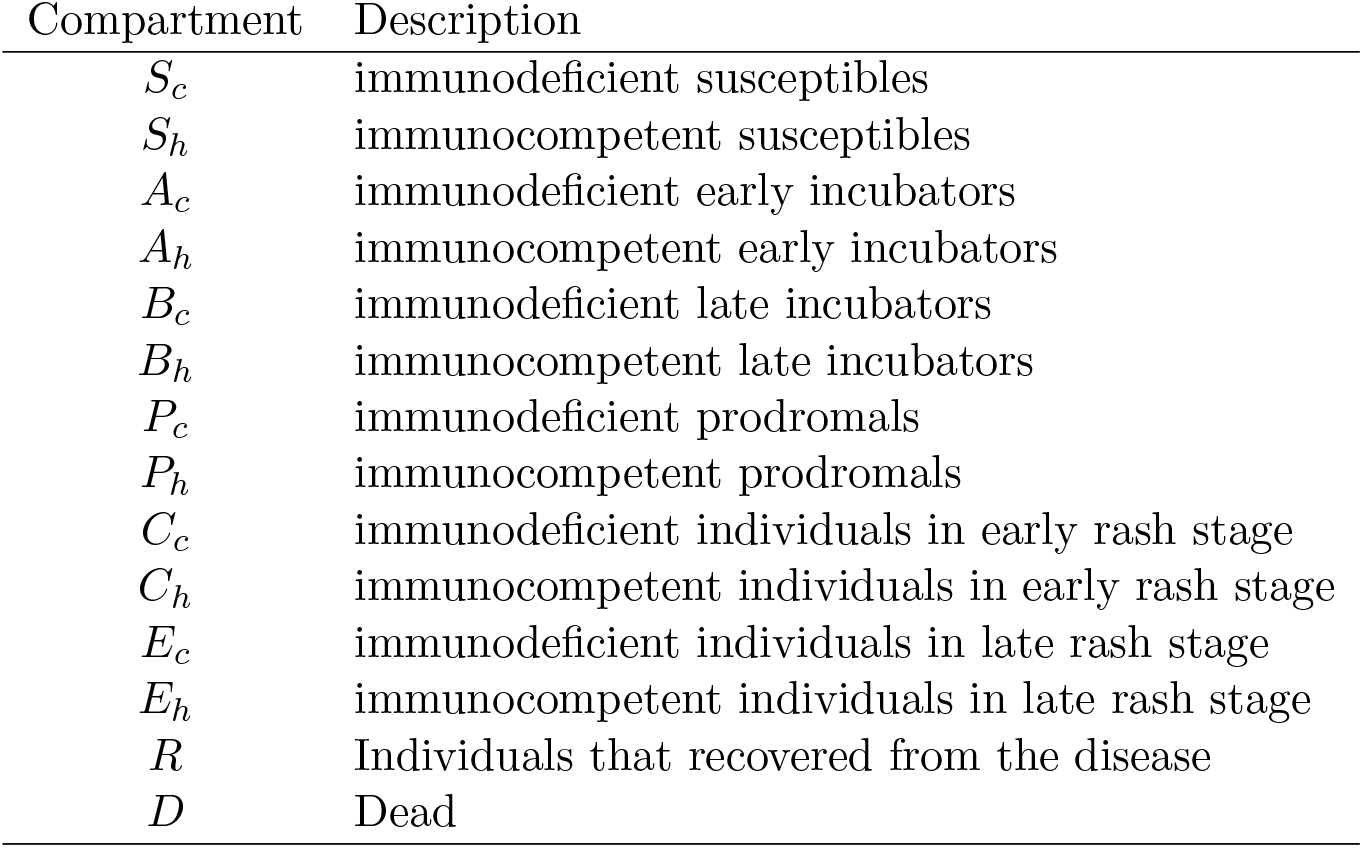
Relevant abbreviations (no countermeasure scenario)

| Compartment | Description |
| --- | --- |
| $S_c$ | immunodeficient susceptibles |
| $S_h$ | immunocompetent susceptibles |
| $A_c$ | immunodeficient early incubators |
| $A_h$ | immunocompetent early incubators |
| $B_c$ | immunodeficient late incubators |
| $B_h$ | immunocompetent late incubators |
| $P_c$ | immunodeficient prodromals |
| $P_h$ | immunocompetent prodromals |
| $C_c$ | immunodeficient individuals in early rash stage |
| $C_h$ | immunocompetent individuals in early rash stage |
| $E_c$ | immunodeficient individuals in late rash stage |
| $E_h$ | immunocompetent individuals in late rash stage |
| $R$ | Individuals that recovered from the disease |
| $D$ | Dead |

We model the evolution of the states as per a set of epidemiological differential equations, which we call *clustered epidemiological differential equations* or the CEDE. Let 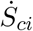 be the derivative of *S*_*ci*_(*t*) with respect to time *t* (i.e., the rate of change with respect to *t*). 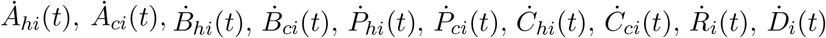 may be defined similarly. The parameters of the CEDE have been summarized and defined in Tables 7 and 8 next.

The CEDE follows:

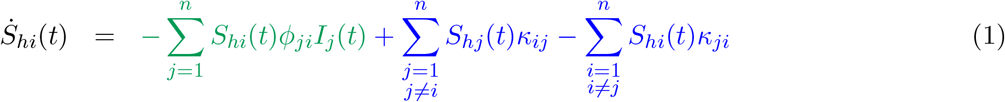

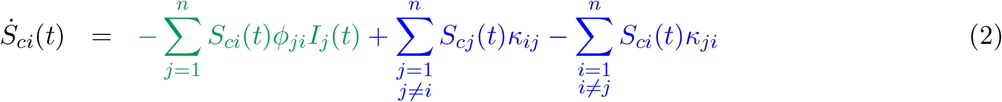

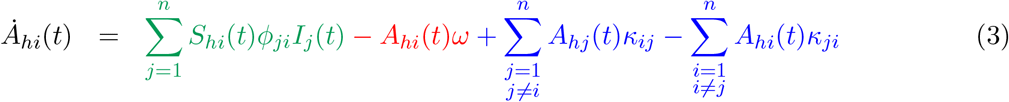

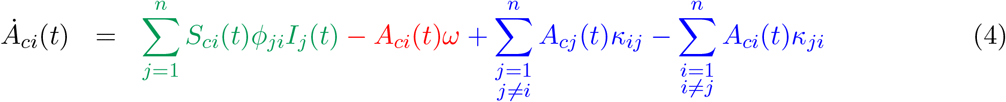

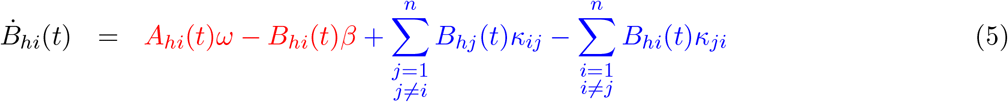

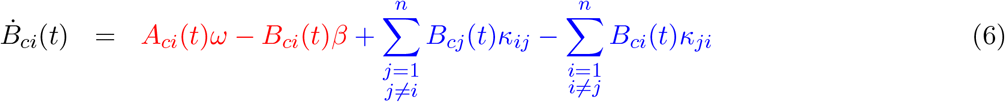

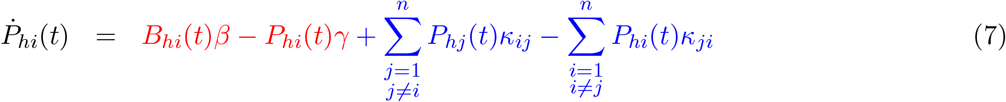

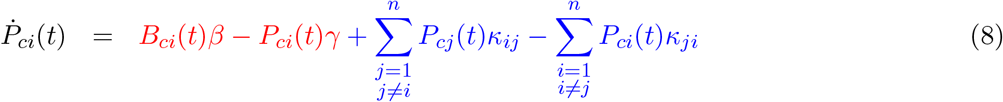

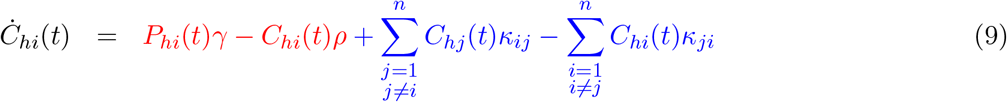

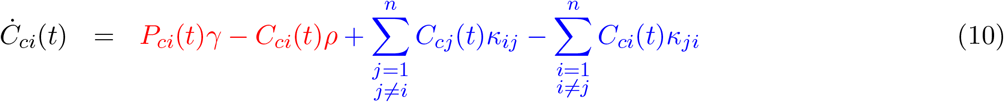

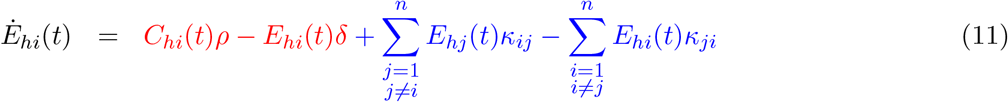

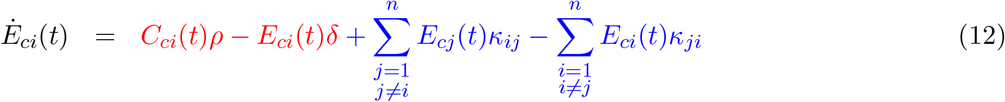

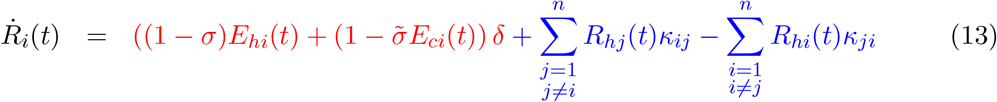

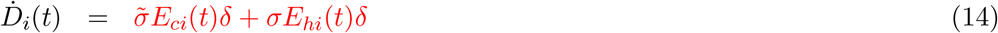

First note that (1) is similar to (2) as transitions are similar for the immunocompetent and immunodeficient individuals. The same observation may be made for ((3), (4)), ((5), (6)), ((7), (8)), ((9), (10)), ((11), (12)). So, we only explain (1), (3), (5), (7), (9), (11), as also (13), (14).

The first terms in (1), (3), (the terms in green color) are quadratic terms which represent the spread of the disease to the susceptibles due to interaction with the infectious individuals and the subsequent transformation of the susceptibles to early incubators - refer to the yellow arrows in Fig 1. Note that quadratic terms represent interactional changes as those changes involve interaction between two individuals (as is typical with epidemiological modeling starting from the classical Kermack–McKendrick work [22]). Since the spread of the disease reduces the number of susceptibles and increases the number of early incubators, the first term in (1) has a positive sign and that in (3) has a negative sign. The parameter *ϕ*_*i,j*_ represents the rate of spread and has been defined in the methods section.

The last two terms in (1), (3), (5), (7), (9), (11), (13) (the terms in blue) are linear terms representing mobility of individuals across clusters. The positive and negative terms respectively represent inflow into and outflow from the cluster. The dead individuals do not move, so we do not see such terms in (14). The mobility rates have been defined in the methods section.

All other terms in (3), (5), (7), (9), (11), (13), and (14) (the terms in red) represent the natural progression of the disease - refer to the yellow arrows in Fig 1. The positive (negative, respectively) terms correspond to arrows directed towards (away, respectively) from the corresponding state. For example the second term in (3) and the first term in (5) represent the natural progression from early incubation to late incubation. The first term in (13) represents the recovery of immunocompetent and immunodeficient individuals from the late rash stage.

Finally, the solution of the CEDE provides the spatio-temporal distribution for the spread of the disease, namely the fraction of individuals who (1) are dead and are in cluster *i* at time *t* (*D*_*i*_(*t*)), (2) have recovered and are in cluster *i* at time *t* (*R*_*i*_(*t*)), (3) are susceptible and are in cluster *i* at time *t* ((*S*_*ci*_ + *S*_*hi*_)(*t*)), (4) are infected and are in cluster *i* at time *t* ((*P*_*ci*_ + *P*_*hi*_ + *C*_*ci*_ + *C*_*hi*_ + *E*_*ci*_ + *E*_*hi*_)(*t*)), (5) are contagious and are in cluster *i* at time *t* ((*S*_*ci*_ + *S*_*hi*_ + *A*_*ci*_ + *A*_*hi*_ + *B*_*ci*_ + *B*_*hi*_ + *P*_*ci*_ + *P*_*hi*_ + *C*_*ci*_ + *C*_*hi*_ + *E*_*ci*_ + *E*_*hi*_)(*t*)).

### A.2 Drug only

Comparing the state transitions for the “no countermeasure” and the “drug only” scenarios, as depicted in Figs 1 and 2, we notice that the latter varies from the former in that it has one additional state called *Preempted* or *Q*. Thus, the CEDE for the drug only scenario can be obtained from (1) to (14) by adding an equation that models the transition to this state and modifying (1) to (14) to reflect this transition. Let *Q*(*t*) be the fraction of individuals who have been preempted after receiving the drug. Then, (15) represents the evolution of *Q*(*t*).

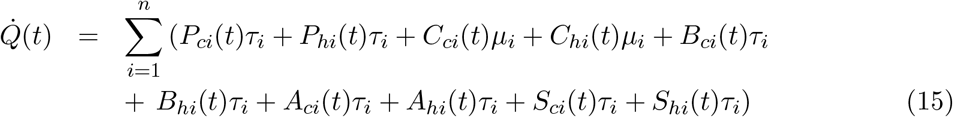

Here *τ*_*i*_ is the reciprocal of the expected time required for an individual to receive the drug in cluster *i* times the probability that the drug preempts the disease given that the individual is yet to reach the rash stage. Similarly, *µ*_*i*_ is the reciprocal of the expected time required for patients in cluster *i* to receive the drug times the preemption probability in the rash stage. The terms *τ*_*i*_*P*_*ci*_(*t*), *τ*_*i*_*P*_*hi*_(*t*), *µ*_*i*_*C*_*ci*_(*t*), *µ*_*i*_*C*_*hi*_(*t*), *τ*_*i*_*A*_*ci*_(*t*), *τ*_*i*_*A*_*hi*_(*t*), *τ*_*i*_*B*_*ci*_(*t*), *τ*_*i*_*B*_*hi*_(*t*), *τ*_*i*_*S*_*ci*_(*t*), *τ*_*i*_*S*_*hi*_(*t*), represent the preemption of the individuals in cluster *i* in the respective states. Due to these preemptions, (1) through (13) will respectively be decremented by *S*_*hi*_*τ*_*i*_, *S*_*ci*_*τ*_*i*_, *A*_*hi*_*τ*_*i*_, *A*_*ci*_*τ*_*i*_, *B*_*hi*_*τ*_*i*_, *B*_*ci*_*τ*_*i*_, *P*_*hi*_*τ*_*i*_, *P*_*ci*_*τ*_*i*_, *C*_*hi*_*µ*_*i*_, and *C*_*ci*_*µ*_*i*_.

The CEDE above can cater to a range of policies for administering of the drug through appropriate choice of the *τ*_*i*_, and *µ*_*i*_. A policy choice may for example be to administer the drug at a higher rate in certain clusters, either because of their centrality and thereby the prospect of infectious individuals therein infecting others at a higher rate, or because the number of infected individuals is high there. Such policies may be represented by selecting a higher value for the corresponding *τ*_*i*_s. A policy choice may be to administer the drug to only those showing certain symptoms, e.g., fever. The *τ*_*i*_s may then be selected based on the stage of the disease - then they would be indexed by the stage as well. Finally, when the drug availability is finite, *τ*_*i*_ and *µ*_*i*_ may be made dependent on time, and as the drug supply is exhausted, the *τ*_*i*_ and *µ*_*i*_ may be chosen to be 0.

Finally, the solution of the CEDE provides the spatio-temporal distribution for the spread of the disease and the impact of the countermeasure (drug). These are captured in the fraction of individuals who (1) are dead and are in cluster *i* at time *t* (*D*_*i*_(*t*)), (2) have recovered and are in cluster *i* at time *t* (*R*_*i*_(*t*)), (3) are preempted (through drug) and are in cluster *i* at time *t* (*Q*_*i*_(*t*)), (4) are susceptible and are in cluster *i* at time *t* ((*S*_*ci*_ + *S*_*hi*_)(*t*)), (5) are infected and are in cluster *i* at time *t* ((*P*_*ci*_ + *P*_*hi*_ + *C*_*ci*_ + *C*_*hi*_ + *E*_*ci*_ + *E*_*hi*_)(*t*)), (6) are contagious and are in cluster *i* at time *t* ((*S*_*ci*_ + *S*_*hi*_ + *A*_*ci*_ + *A*_*hi*_ + *B*_*ci*_ + *B*_*hi*_ + *P*_*ci*_ + *P*_*hi*_ + *C*_*ci*_ + *C*_*hi*_ + *E*_*ci*_ + *E*_*hi*_)(*t*)).

### A.3 Vaccine only

The principal difference between the “Vaccine only” scenario and the earlier scenarios lies in the incorporation of the opinion dynamics. The individuals must now be classified based on their cooperativity, leading to the new states enunciated in Table 9.

**Table 7:** Parameters for the CEDE in the no countermeasure scenario.

| Parameter | Symbols | Some associated terms in (1) - (14) |
| --- | --- | --- |
| Mobility rate | $\kappa_{i,j}$ | $S_{hj}(t)\kappa_{ij}, B_{ci}(t)\kappa_{ji}, E_{hj}(t)\kappa_{ij}, P_{ci}(t)\kappa_{ji}, etc.$ |
| Disease spread rate | $\phi_{i,j}$ | $S_{hi}(t)\phi_{ji}I_j(t), S_{ci}(t)\phi_{ji}I_j(t)$ |
| Opinion spread rate | $\alpha_{i,j}$ | This scenario is not affected by opinion |
| Disease transition rate | $\omega, \beta, \gamma, \rho, \delta$ | $A_{hi}(t)\omega, P_{ci}(t)\beta, C_{hi}(t)\rho, R_{ci}(t)\delta, etc.$ |

**Table 8:** The disease transition rate parameters.

| Parameters | Description of parameters |
| --- | --- |
| $\frac{1}{\omega}$ | Expected time an individual is in early incubation |
| $\frac{1}{\beta}$ | Expected time an individual is in late incubation |
| $\frac{1}{\gamma}$ | Expected time an individual is in prodrome stage |
| $\frac{1}{\rho}$ | Expected time an individual is in early rash stage |
| $\frac{1}{\delta}$ | Expected time an individual is in late rash stage |
| $\sigma$ | Probability that an immunocompetent individual at late rash stage dies |
| $\tilde{\sigma}$ | Probability that an immunodeficient individual at late rash stage dies |

**Table 9:** Relevant abbreviations (vaccine only scenario)

| Compartment | Description |
| --- | --- |
| $S_{ac}$ | immunodeficient cooperative susceptibles |
| $S_{bc}$ | immunodeficient non-cooperative susceptibles |
| $S_{ah}$ | immunocompetent cooperative susceptibles |
| $S_{bh}$ | immunocompetent non-cooperative susceptibles |
| $A_{ac}$ | immunodeficient cooperative early incubators |
| $A_{bc}$ | immunodeficient non-cooperative early incubators |
| $A_{ah}$ | immunocompetent cooperative early incubators |
| $A_{bh}$ | immunocompetent non-cooperative early incubators |
| $B_{ac}$ | immunodeficient cooperative late incubators |
| $B_{bc}$ | immunodeficient non-cooperative late incubators |
| $B_{ah}$ | immunocompetent cooperative late incubators |
| $B_{bh}$ | immunocompetent non-cooperative late incubators |
| $P_{ac}$ | immunodeficient cooperative prodromals |
| $P_{bc}$ | immunodeficient non-cooperative prodromals |
| $P_{ah}$ | immunocompetent cooperative prodromals |
| $P_{bh}$ | immunocompetent non-cooperative prodromals |
| $C_{ac}$ | immunodeficient cooperative individuals in early rash stage |
| $C_{bc}$ | immunodeficient non-cooperative individuals in early rash stage |
| $C_{ah}$ | immunocompetent cooperative individuals in early rash stage |
| $C_{bh}$ | immunocompetent non-cooperative individuals in early rash stage |
| $E_{ac}$ | immunodeficient cooperative individuals in late rash stage |
| $E_{bc}$ | immunodeficient non-cooperative individuals in late rash stage |
| $E_{ah}$ | immunocompetent cooperative individuals in late rash stage |
| $E_{bh}$ | immunocompetent non-cooperative individuals in late rash stage |
| $R_a$ | Cooperative individuals that recovered from the disease |
| $R_b$ | Non-cooperative individuals that recovered from the disease |
| $Q$ | Vaccinated (immunized) |
| $D$ | Dead |

To consider the impact of space, we further classify individuals in the states given in Table 9 based on the cluster they inhabit. Thus, *S*_*ahi*_(*t*) (*S*_*bci*_(*t*), respectively) is the fraction of immunocompetent cooperative (immunodeficient non-cooperative, respectively) susceptibles who are in cluster *i* at time *t*. As before, *I*_*i*_(*t*) is the fraction of the *infectious* individuals in cluster *i* at time *t*. Let *X*_*i*_(*t*) be the fraction of individuals who are cooperative and are in cluster *i* at time *t*. Thus,

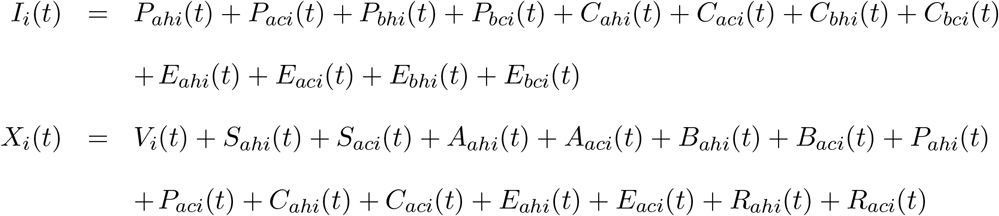

The spread of opinions is modeled in the CEDE through quadratic terms (the terms in orange) representing opinion exchange interactions between the non-cooperative and the cooperative individuals, just as the spread of disease is modeled through quadratic terms (the terms in green) representing physical contact based interactions between the susceptible and infectious individuals. For simplicity, in this subsection, we present the CEDE only for the scenario that during opinion exchanges the cooperatives persuade the noncooperatives to become cooperatives.

The immunocompetent cooperative susceptibles (respectively early incubators) who are in cluster *i* are preempted (through vaccination) at the rate *θ*_*i*_ (*π*_*i*_ respectively). The preemption rate is the product of the reciprocal of the expected delay in developing immunity and the probability that the vaccine provides immunity at the stage of the disease the individual is in. The expected delay is the sum of the expected delays in 1) delivering the vaccine (e.g., delay incurred in getting the health worker’s appointment) and 2) developing immunity after the vaccine is administered. After these individuals are vaccinated and develop immunity, they transition to preempted state *Q* (the terms in black in the CEDE below); this process is denoted by the yellow arrows in Fig 3. The fraction of individuals who are vaccinated and are in cluster *i* at time *t* is *Q*_*i*_(*t*).

The resulting CEDE, for the evolution of the states of the immunocompetent individuals, is shown in (16) - (31) with the coefficients defined in Tables 7 and 8 and additional parameters defined in Table 10.

**Table 10:**
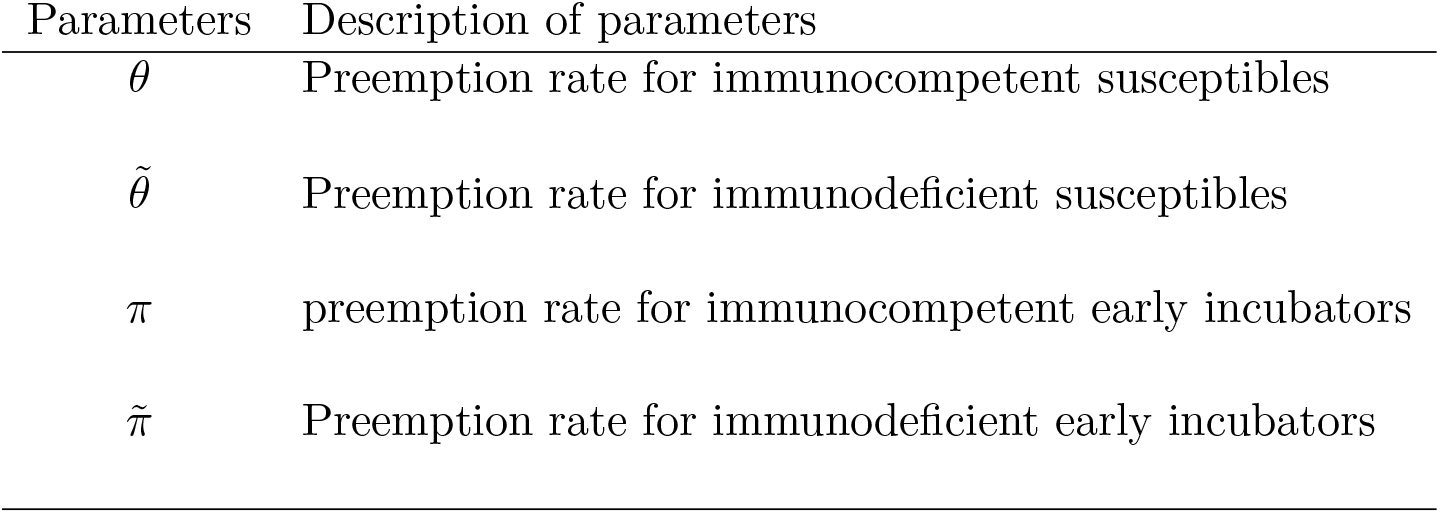
Parameters description (vaccine only scenario)

| Parameters | Description of parameters |
| --- | --- |
| $\theta$ | Preemption rate for immunocompetent susceptibles |
| $\tilde{\theta}$ | Preemption rate for immunodeficient susceptibles |
| $\pi$ | preemption rate for immunocompetent early incubators |
| $\tilde{\pi}$ | Preemption rate for immunodeficient early incubators |

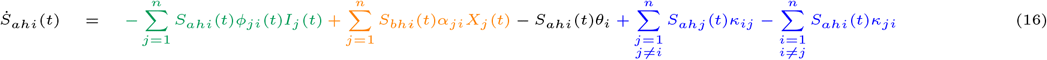

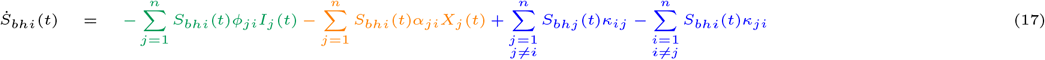

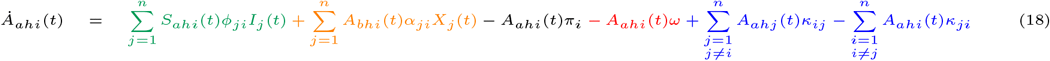

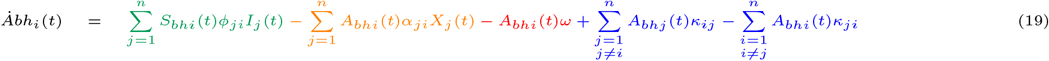

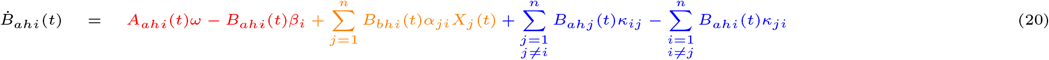

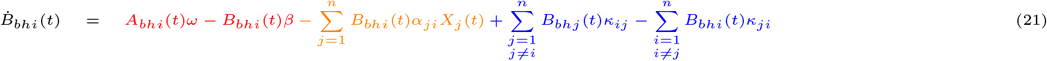

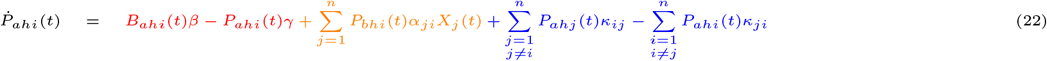

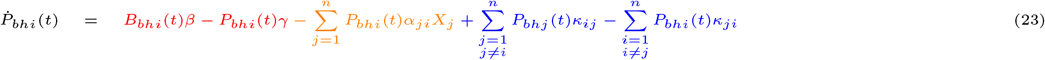

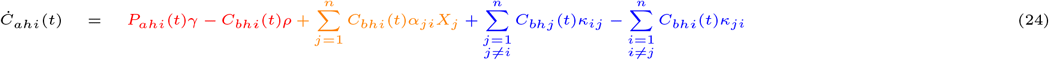

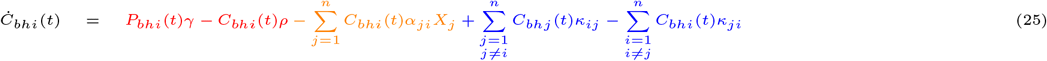

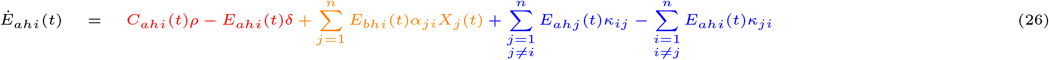

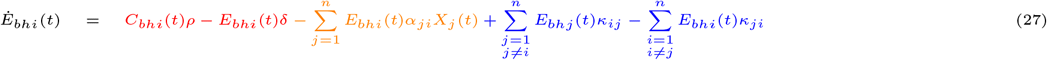

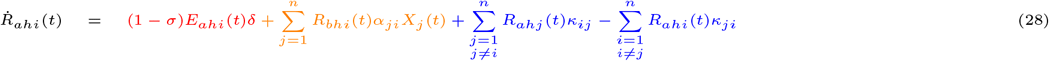

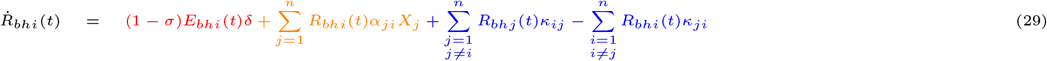

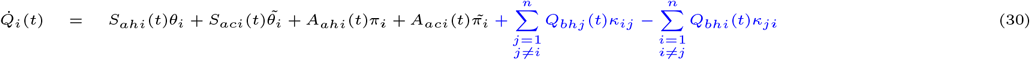

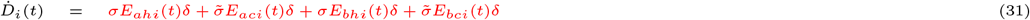

The differential equations for the evolution of the states of the immunodeficient individuals is identical to those of the immunocompetent individuals, as seen in equations (1) to (12) in Appendix A.1. Specifically, the equation for 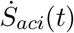 may be obtained from equation (16) for 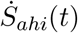, as the equation (2) for 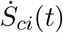 followed from equation (1) for 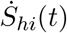. The only differences in the equations for the immunodeficient individuals are that 1) they have lower preemption rates than the immunocompetent ones as they can avail of different vaccines that act slower and 2) those in late rash stage die while the corresponding immunocompetent ones can recover. Specifically, the immunodeficient cooperative susceptibles (respectively early incubators) that are in cluster *i* develop immunity at rate 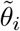 (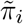 respectively). The preemption rates are defined as for the immunocompetent ones, but have lower value as the immunodeficient ones can only receive vaccines that provide immunity after multiple shots administered after specified gaps. Thus, in general, 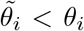 and 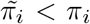. Note that the recovery and death rates for the immunodeficient and the immunocompetent ones are as in Table 8.

We now explain the terms in (16) - (31). The second terms in (16) through (29) (the terms in orange) are quadratic terms which represent the transformation of non-cooperative individuals to cooperative ones through exchange of opinion with cooperative individuals - refer to the black arrows in Fig 3. Since the change of opinion increases (respectively decreases) the cooperatives (respectively noncooperatives), the second terms in the equations for the cooperatives (e.g., (16)) are positive and the second terms for the non-cooperatives (e.g., (17)) are negative.

The terms *θ*_*i*_*S*_*ahi*_(*t*) and *π*_*i*_*A*_*ahi*_(*t*) (the terms in black) in (30) represent the vaccination of immunocompetent cooperative susceptibles and early incubators. Vaccination reduces the number of susceptibles and early incubators by transforming them to the vaccinated state. Thus, these terms are subtracted from (16) and (18) respectively and added to (30). The second and fourth terms of equation (30) correspond to the immunization of immunodeficient cooperative susceptibles and early incubators.

The rest of the terms represent the phenomena modeled in equations (1) to (14) in Appendix A.1. Specifically, the first terms in (16), and (18) (the terms in green) are quadratic terms which represent the spread of the disease to the susceptibles due to interaction with the infectious individuals and the subsequent transformation of the susceptibles to early incubators - refer to the yellow arrows in Fig 3. The disease dynamics is similar for the cooperatives and non-cooperatives. The last two terms in (16), (18), (20), (22), (24), (26), (28) (the terms in blue) are linear terms representing mobility of individuals across clusters. The dead individuals do not move, so we do not see such terms in (31). All other terms in (16), (18), (20), (22), (24), (26) (the terms in red) represent the natural progression of the disease - refer to the blue arrows in Fig 3.

Different types of vaccines and immunization policies may be captured through appropriate choices of the values of 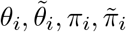. A policy choice may for example be to administer the vaccine at a higher rate in certain clusters. Such policies may be represented by selecting a higher value for the corresponding parameters. Some vaccines may act faster than others leading again to higher values for the above parameters. Finally, when the availability of the vaccine is finite, the values of these parameters may be made dependent on time, and as the supply of the vaccine is exhausted, the parameters may be chosen to be 0.

Finally, the solution of the CEDE provides the spatio-temporal distribution for the spread of the disease and opinion and the impact of the countermeasure (vaccine). These are captured in the fraction of individuals who (1) are dead and are in cluster *i* at time *t* (*D*_*i*_(*t*)), (2) have recovered and are in cluster *i* at time *t* ((*R*_*ahi*_ + *R*_*bhi*_)(*t*)), (3) are preempted (that is, vaccinated) and are in cluster *i* at time *t* (*Q*_*i*_(*t*)), (4) are susceptible and are in cluster *i* at time *t* ((*S*_*aci*_ + *S*_*ahi*_ + *S*_*bci*_ + *S*_*bhi*_)(*t*)), (5) are infected and are in cluster *i* at time *t* ((*P*_*aci*_ + *P*_*ahi*_ + *P*_*bci*_ + *P*_*bhi*_ + *C*_*aci*_ + *C*_*ahi*_ + *C*_*bci*_ + *C*_*bhi*_ + *E*_*aci*_ + *E*_*ahi*_ + *E*_*bci*_ + *E*_*bhi*_)(*t*)), (6) are contagious and are in cluster *i* at time *t* ((*S*_*aci*_ + *S*_*ahi*_ + *S*_*bci*_ + *S*_*bhi*_ + *A*_*aci*_ + *A*_*ahi*_ + *A*_*bci*_ + *A*_*bhi*_ + *B*_*aci*_ + *B*_*ahi*_ + *Bbci* + *Bbhi* + *Paci* + *Pahi* + *Pbci* + *Pbhi* + *Caci* + *Cahi* + *Cbci* + *Cbhi* + *Eaci* + *Eahi* + *Ebci* + *Ebhi*)(*t*)).

### A.4 Both drug and vaccine

In this case, individuals may be preempted through drug or vaccine. Thus, those preempted may either be cooperative or non-cooperatives. Thus, we classify the preempted individuals into cooperatives, *Q*_*a*_ and non-cooperatives, *Q*_*b*_ (Fig 4). The cooperatives among the susceptibles and early incubators can transition to the state *Q*_*a*_ after developing immunity through drug or vaccine. Transition from the rest of the states (other than recovered or dead) to *Q*_*a*_ or *Q*_*b*_ occurs through reception of the drug. *Q*_*a*_, *Q*_*b*_ needs to be indexed by the clusters the individuals inhabit.

We use the parameters as defined in Appendix A.3 with some of the modifications described in Table 11. Thus, the CEDE for both drug and vaccine scenario can be obtained from (16) to (29) by adding two equations, (32) and (33) below, that model the transitions to *Q*_*a*_, *Q*_*b*_ and modifying (16) to (29) to reflect this transition and the distinction between *Q*_*a*_, *Q*_*b*_.

**Table 11:** Parameters description (both drug and vaccine)

| Parameters | Description of parameters |
| --- | --- |
| $\Theta$ | Rate of preemption for immunocompetent cooperative susceptibles through the reception of the drug or the vaccine |
| $\tilde{\Theta}$ | Rate of preemption for immunodeficient cooperative susceptibles through the reception of the drug or the vaccine |
| $\Pi$ | Rate of preemption for immunocompetent cooperative early incubators through the reception of the drug or the vaccine. |
| $\tilde{\Pi}$ | Rate of preemption for immunodeficient early incubators through the reception of the drug or the vaccine. |
| $\bar{\Theta}$ | Rate of preemption for immunocompetent non-cooperative susceptibles through the reception of the drug or the vaccine |
| $\hat{\Theta}$ | Rate of preemption for immunodeficient non-cooperative susceptibles through the reception of the drug or the vaccine |
| $\bar{\Pi}$ | Rate of preemption for immunocompetent non-cooperative early incubators through the reception of the drug or the vaccine. |
| $\hat{\Pi}$ | Rate of preemption for immunodeficient non-cooperative early incubators through the reception of the drug or the vaccine. |
| $\tau$ | Rate of preemption for prodromals and non-cooperatives among susceptibles through reception of drugs |
| $\mu$ | Rate of preemption for patients with rash through reception of drugs. |

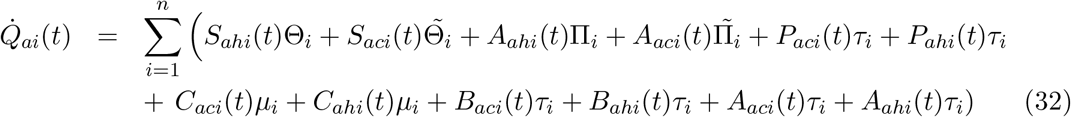

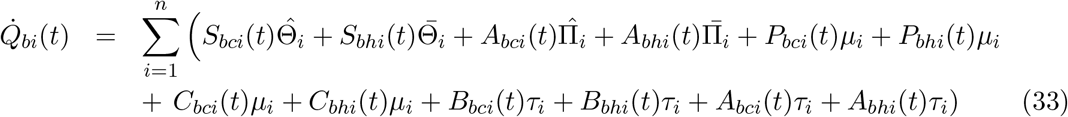

We now describe the modifications in (16) to (29). The expression for *X*_*i*_(*t*) in Appendix (A.3) need to have *Q*_*ai*_(*t*) instead of *Q*_*i*_(*t*). The terms 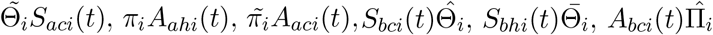, and 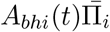 represent the preemption of individuals by either vaccine or drugs for individuals in cluster *i* in their respective states. In addition, *τ*_*i*_*P*_*ahi*_(*t*), *τ*_*i*_*P*_*aci*_(*t*), *τ*_*i*_*P*_*bhi*_(*t*), *τ*_*i*_*P*_*bci*_(*t*), *µ*_*i*_*C*_*ahi*_(*t*), *µ*_*i*_*C*_*aci*_(*t*), *µ*_*i*_*C*_*bhi*_(*t*), *µ*_*i*_*C*_*bci*_(*t*), *µ*_*i*_*E*_*ahi*_(*t*), *µ*_*i*_*E*_*aci*_(*t*), *τ*_*i*_*B*_*ahi*_(*t*), *τ*_*i*_*B*_*aci*_(*t*), *τ*_*i*_*B*_*bhi*_(*t*), and *τ*_*i*_*B*_*bci*_(*t*) represent preemption by drug only for individuals in cluster *I* in the respective states. Due to these preemptions, (16) through (27) will respectively be decremented by 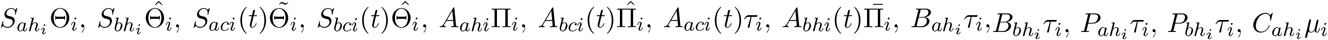, and 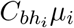.

Here the suffices *a, b* represent the two opinions. Those with suffix *a* convert those with suffix *b*. To capture the scenario that cooperatives convert the non-cooperatives, we consider those with suffix *a* as cooperative, and those with suffix *b* as non-cooperative. To capture the scenario that non-cooperatives convert the cooperatives, we reverse the designations, that is, those with suffix *a* are non-cooperatives, and those with suffix *b* are cooperatives. Note that functionally the difference between the cooperatives and non-cooperatives is that the former opt for vaccination while the latter does not. The vaccinations can be effective only if the individuals are susceptible or in early incubation stage. Thus, in these states, the cooperatives can be preempted through either drugs or vaccines, while the non-cooperatives can only be preempted through drugs (that is, if they are eligible for drugs depending on the administering policies). Thus, the preemption rates of the cooperatives are different from the non-cooperatives in these stages, and that is the only functional difference. Thus, interchanging the preemption rates of the states with suffix *a* with those of the corresponding ones with suffix *b* will switch their roles as cooperatives and non-cooperatives. We describe how the preemption rates are chosen in Section B of Appendix.

Finally, various policy choices for countermeasure application may be incorporated through appropriate selection of the parameters in Table 11, similar to the connections made in the concluding paragraphs of Appendices A.2 and A.3. In addition, the solution of the CEDE provides the spatio-temporal distribution for the spread of the disease and opinion and the impact of the countermeasure (vaccine and drug). These are captured in fraction of individuals who (1) are dead and are in cluster *i* at time *t* (*D*_*i*_(*t*)), (2) have recovered and are in cluster *i* at time *t* ((*R*_*ahi*_ + *R*_*bhi*_)(*t*)), (3) are preempted (that is, vaccinated or administered drugs) and are in cluster *i* at time *t* ((*Q*_*ai*_ + *Q*_*bi*_)(*t*)), (4) are susceptible and are in cluster *i* at time *t* ((*S*_*aci*_ + *S*_*ahi*_ + *S*_*bci*_ + *S*_*bhi*_)(*t*)), (5) are infected and are in cluster *i* at time *t* ((*P*_*aci*_ + *P*_*ahi*_ + *P*_*bci*_ + *P*_*bhi*_ + *C*_*aci*_ + *C*_*ahi*_ + *C*_*bci*_ + *C*_*bhi*_ + *E*_*aci*_ + *E*_*ahi*_ + *E*_*bci*_ + *E*_*bhi*_)(*t*)), (6) are contagious and are in cluster *i* at time *t* ((*S*_*aci*_ + *S*_*ahi*_ + *S*_*bci*_ + *S*_*bhi*_ + *A*_*aci*_ + *A*_*ahi*_ + *A*_*bci*_ + *A*_*bhi*_ + *B*_*aci*_ + *B*_*ahi*_ + *B*_*bci*_ + *B*_*bhi*_ + *P*_*aci*_ + *P*_*ahi*_ + *P*_*bci*_ + *P*_*bhi*_ + *C*_*aci*_ + *C*_*ahi*_ + *C*_*bci*_ + *C*_*bhi*_ + *E*_*aci*_ + *E*_*ahi*_ + *E*_*bci*_ + *E*_*bhi*_)(*t*)).

## B Parameter estimation

### B.1 Estimation of parameters used in all the scenarios

We first estimate the parameters that occur in all four scenarios ((1) no countermeasure (2) drug only (3) vaccine only (4) both drug and vaccine) we have considered so far, namely the disease progression parameters. These parameters have been described in Table 8. Typically, the early incubation phase for smallpox lasts for 7 days and the late incubation phase for 5 days [14]. Assuming that each phase has an exponentially distributed random duration, 1*/ω*, 1*/β* are the expected durations in these two phases. Thus, *ω* = 1*/*7, *β* = 1*/*5. Typically a patient is in prodromal phase for 3 days [14, 15]. Thus, as for the incubation phases, *γ* = 1*/*3. Typically, the early and late rash periods last for 3 and 7 days respectively [14]. Therefore, as before, *ρ* = 1*/*3, *δ* = 1*/*7. On average, 30% of the infected immunocompetent individuals die of the disease [14, 15]. Thus, *σ* = 0.3. According to [13], immunodeficient individuals are not expected to recover from smallpox. Thus, 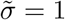.

We now compute the disease spread rate described in Table 7. We assume *ϕ*_*i,j*_ = 0 if *i* ≠ *j*, that is, there is no direct physical contact and therefore no direct spread of disease between people inhabiting different clusters. The *basic reproductive number, R*_0_, is defined as the average number of secondary infections caused by a single typical infected individual among a completely susceptible population. [11, 14] estimated *R*_0_ for smallpox to be 6.9. We estimate *ϕ*_*i,i*_ to obtain *R*_0_ = 6.9. Following the methodology in [20, 32], we get *ϕ*_*i,i*_ = 0.00173.

Note that the infection indirectly spreads between clusters due to mobility of individuals across clusters. Mobility rate is described in the methods section and Table 7. We do not assume any specific value of the mobility rate between clusters but vary it across a range of values as mobility rates heavily depend on the scenario under consideration.

### B.2 Estimation of parameters related to delivery of drugs

We now estimate the parameters related to the preemption of infected individuals by drugs (refer to Appendix A.2), these will also be used to compute the parameters in both drug and vaccine scenarios. First, note that the stage of the disease in which an individual can be administered a drug depends on the drug administering policy. We assume that in a state in which an individual is allowed to receive a drug, he needs to wait for an exponentially distributed random time after which he is preempted, that is, he is administered the drug and develops the ability to thwart the disease. Let *ε* be the parameter of this exponential process. We also assumed that the wait time for different individuals is independent.

There are *n* individuals in all. The first individual who is preempted waits for the minimum time among *n*. From the theory of exponential distributions, the minimum time is exponentially distributed with parameter *nε*, and has expectation 1*/nε*. The second person to be preempted has the minimum additional wait time among the remaining *n* − 1. Similarly, this additional wait time is exponentially distributed with parameter (*n* − 1)*ε* and expectation 1*/*(*n* − 1)*ε*. Similarly, the additional wait time for the third person (after the second person is preempted) is exponential with parameter (*n* − 2)*ε*, and has expectation 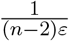. This continues until the last person whose expected additional wait time is 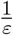. Therefore, the total expected time for n individuals to be preempted is,

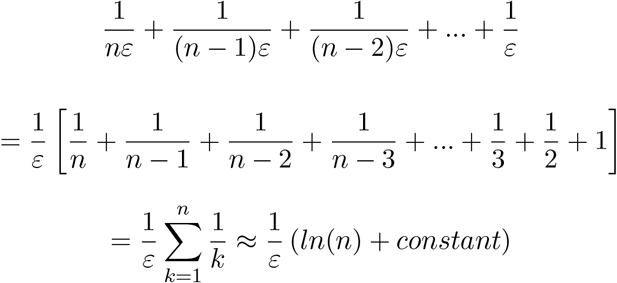

The last expression follows from the Harmonic series (number) approximation. We obtain *ε* by equating this expression to the total wait times required to develop immunity as reported in the literature for cities. [14] and [33] state that the CDC’s Cities Readiness Initiative sets as a goal that cities should be able to distribute antibiotics to their entire population within 48 hours. However, it is uncertain how many large cities with a population similar to our model are prepared to meet this goal. Therefore, similar to [14], we consider a mass antiviral distribution timetable of 4 days. We assumed that an individual can thwart the disease 2 days on average after he starts receiving the drug. We also assume that the constant in the Harmonic number approximation is negligible. Therefore, *ε* = *ln* (*n*) */*(4 + 2). Considering *n* = 10^7^, we have *ε* = 2.6863.

According to [14], drug treatment before the onset of rash offers 99% protection against mortality, and drug treatment 3 days post-onset of rash offers 80% protection against mortality. Thus, we estimate the preemption rate for susceptibles, incubators, and prodromals as *τ* = *ε ×* 0.99 = 2.6594 (if the drug administering policy administers the drug to them), and the preemption rate for those in the early rash stage as *µ* = *ε* * 0.8 = 2.1490. We assume that the drug is ineffective in the late rash stage.

All policies for administering drugs use *µ* = 2.1490 as computed above. But *τ* will be 0 for some states in which the drug is not administered depending on the policy under consideration. Note that policy 1 administers drugs to only the patients with rashes. Thus, under policy 1, *τ* = 0. In policy 2, once the number of cases in a given cluster exceeds a certain threshold, drug is administered to anyone with fever (prodrome) or rashes, and *τ* = 2.6594 for prodromals; otherwise, *τ* = 0 as in policy 1. In policy 3, drugs are administered to everyone in a given cluster once the number of cases in the cluster exceeds a certain threshold, otherwise only those with rash receive the drug. Thus, once the number of cases in a cluster exceed a certain threshold, *τ* = 2.6594, otherwise *τ* = 0.

### B.3 Estimating the parameters pertaining to immunization

We now consider the additional parameters in the vaccine only scenario above and beyond those estimated in Appendix B.1. Refer to Table 10. According, to [14, 16, 20], it would take approximately 10 days on average for cooperative individuals to get their shots in case of mass vaccination against an epidemic in a large U.S. city. The immunocompetent individuals are given the live vaccine (ACAM2000) which provides immediate immunity if taken during the susceptible stage [26]. Finally, according to [14], vaccination within the early incubation stage provides an 80% chance of disease prevention. Now, following the methodology in Appendix B.2, 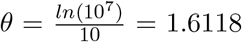 and 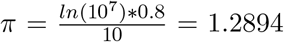 (as in Appendix B.2, we consider that there are *n* = 10^7^ individuals in the city under consideration). Next, we consider immunodeficient individuals. They receive the Modified Vaccinia virus Ankara (MVA) vaccines [11,14] that takes two-shots 30 days apart to provide immunity [17]. Thus 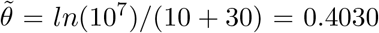, and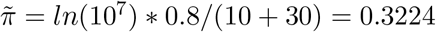.

### B.4 Modifying the estimation for the vaccine and drug scenario

In this scenario, individuals can be preempted through either drug or vaccine. We estimate the parameters in Table 11. We first consider the default case, that is, the cooperatives convert the non-cooperatives, in this, the states with *a* in the suffix are cooperatives and those with *b* in the suffix are the non-cooperatives. Vaccines are effective only when they are administered to individuals in susceptible or early incubation stage. Thus, preemption in later stages can only be through drugs. Referring to Table 11, 1*/τ* and 1*/µ* can therefore be directly obtained from the estimation strategy in Appendix B.2, i.e., *τ* = 2.6594 and *µ* = 2.1490.

The estimation of other parameters in Table 11 will depend on the policy for administering the drugs. Note that in policy 1, drugs are administered to only patients with rash. Thus, a susceptible or early incubator can be preempted only when he receives the vaccine. Thus 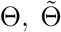Π, and 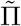 can be obtained directly from the respective values of 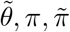 estimated in Appendix B.3. Specifically 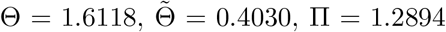 and 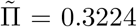. And, since the non-cooperatives do not receive vaccine, 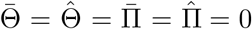.

In policy 2, once the number of cases in a given cluster exceeds a certain threshold, drug is administered to anyone with fever (prodrome) or rash. Thus, a susceptible or early incubator can be preempted only when he receives the vaccine, and Θ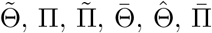, and 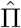 can be estimated identical to the previous paragraph. The drug delivery parameter in the prodrome stage can be estimated as in the first paragraph, i.e., *τ* = 2.6594.

In policy 3, drugs are administered to everyone in a given cluster once the number of cases in the cluster exceeds a certain threshold. Thus, in this case, susceptibles and early incubators may either be preempted through drugs or vaccines. We assume that any such immunocompetent individual is preempted through vaccination after an exponentially distributed wait time with parameter *θ*, and is preempted through drug after an exponentially distributed wait time with parameter *τ*. Thus, he is preempted after a minimum of the two wait times (if he receives the drug earlier than the vaccine he will be preempted through drug and vice versa). When the two exponential processes are independent, their minimum is an exponential process with parameter Θ, where Θ = *θ* + *τ*. Now from the estimations in Appendix A.4 and first 4 rows of Table 11, Θ = 1.6118 + 2.6594 = 4.2712. For an immunodeficient individual the parameter for the vaccine is 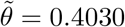 (from Appendix B.3). Thus, 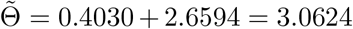. Similarly, Θ = *π* + *τ* = 1.2894 + 2.6594 = 3.9488, and the 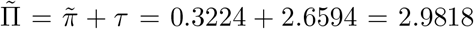. For late incubation, preemption can happen only through drug delivery, and the preemption rate as estimated in the first paragraph is *τ* = 2.6594. The non-cooperative susceptibles and early incubators can only be preempted through drugs. Thus 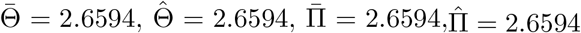.

Finally, we consider the case that the non-cooperatives convert the cooperatives. As explained in Appendix A.4, here the states with suffix *a* are the non-cooperatives and those with suffix *b* are the cooperatives. Since the functional difference between these two are in the pre-emption rates, the preemption rates of the states with suffix *a* need to be interchanged with the corresponding ones with suffix *b*. Note that only the susceptibles and early incubators can receive vaccines, all the rest are preempted only through drugs and therefore their preemption rates are the same regardless of whether they are cooperatives or non-cooperatives. Thus, only the preemption rates of the cooperative susceptibles (respectively, early incubators) need to be interchanged with the corresponding non-cooperative susceptibles (respectively, early incubators). That is, values of Θ, 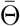 need to be exchanged. Similarly, values of 1) 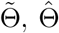, 2) Π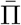, 3) 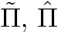 need to be exchanged.

